# Using machine learning to predict COVID-19 infection and severity risk among 4,510 aged adults: a UK Biobank cohort study

**DOI:** 10.1101/2020.06.09.20127092

**Authors:** Auriel A. Willette, Sara A. Willette, Qian Wang, Colleen Pappas, Brandon S. Klinedinst, Scott Le, Brittany Larsen, Amy Pollpeter, Tianqi Li, Nicole Brenner, Tim Waterboer

## Abstract

**Background:** Many risk factors have emerged for novel 2019 coronavirus disease (COVID-19). It is relatively unknown how these factors collectively predict COVID-19 infection risk, as well as risk for a severe infection (i.e., hospitalization).

**Methods:** Among aged adults (69.3 ± 8.6 years) in UK Biobank, COVID-19 data was downloaded for 4,510 participants with 7,539 test cases. We downloaded baseline data from 10-14 years ago, including demographics, biochemistry, body mass, and other factors, as well as antibody titers for 20 common to rare infectious diseases. Permutation-based linear discriminant analysis was used to predict COVID-19 risk and hospitalization risk. Probability and threshold metrics included receiver operating characteristic curves to derive area under the curve (AUC), specificity, sensitivity, and quadratic mean.

**Results:** The “best-fit” model for predicting COVID-19 risk achieved excellent discrimination (AUC=0.969, 95% CI=0.934-1.000). Factors included age, immune markers, lipids, and serology titers to common pathogens like human cytomegalovirus. The hospitalization “best-fit” model was more modest (AUC=0.803, 95% CI=0.663-0.943) and included only serology titers.

**Conclusions:** Accurate risk profiles can be created using standard self-report and biomedical data collected in public health and medical settings. It is also worthwhile to further investigate if prior host immunity predicts current host immunity to COVID-19.

## Introduction

Coronavirus disease 2019 (COVID-19), caused by a novel beta-coronavirus called severe acute respiratory syndrome coronavirus 2 (SARS-CoV-2)^1^, is a worldwide pandemic that continues to severely disrupt the economic, social, and psychological well-being of countless people. Clinical presentation of COVID-19 widely varies, ranging from asymptomatic profiles to mild symptoms like high fever or cough to acute respiratory disease syndrome and death. Given this heterogeneous symptom presentation, as well as difficulties with serology testing, contact tracing, and more recently vaccine administration, it remains important to isolate or maximize safety for adults most at risk for COVID-19 infection and severe disease.

By extension, a large body of research has investigated potential factors that increase COVID-19 infection and disease severity risk. It is well known, for example, that adults aged >65 years are much more likely to be hospitalized or die due to COVID-19. Obesity itself and adverse health behaviors like smoking also increase infection risk and likelihood of hospitalization^2,3^. Several age and obesity-related conditions such as cardiovascular disease, cardiometabolic diseases (e.g., type 2 diabetes), hypertension, and other disease states and syndromes are also of concern^4^. Non-white ethnicity, particularly being black regardless of country of origin, socioeconomic deprivation, and low levels of education even after adjustment for health factors point to less privilege unfortunately conferring risk^5^. Among biological markers, COVID-19 infection or severity has been related to higher C-Reactive Protein and more circulating white blood cells and lower counts of lymphocytes or granulocytes (e.g., monocytes)^6-8^. SARS-CoV-1 has a similar profile except for a relatively normal total white blood cell count^9^.

These studies are invaluable for establishing or validating risk factors to guide clinical decisions and policymaker choices. However, we ultimately need to develop risk profiles derived from these factors to accurately predict who will and will not develop COVID-19, and if a COVID-19 disease course will be mild or presumptively severe (i.e., require hospitalization). Data-driven modelling using machine learning can be used to create robust prediction models based on routinely collected biomedical data like demographics, a complete blood count, and standard medical biochemistry data. Critically, by using non COVID-19 serological data, we may gain insight into the host’s ability to fight COVID-19 by examining antibody titers that detail the host response to past infectious pathogens. This “virome” may affect host innate and adaptive immunity^9,10^. For example, human cytomegalovirus vastly changes the composition of T and B cells^11^, and may induce immune senescence that could account for worse SARS-CoV-2 infection outcomes.

Therefore, our objective was to use classification machine learning to determine how baseline measures, collected 10-14 years ago, could best predict which older adults developed COVID-19. Our second objective was to make similar predictions but for determining if someone positive with COVID-19 had a mild or severe infection. In summary, we achieved > 90% sensitivity and specificity with outstanding diagnostic value (AUC=0.969) for correctly predicting COVID-19 infection based on factors like age, biochemistry and white blood cell markers, and antibody titers to common pathogens like human cytomegalovirus, human herpesvirus 6, and chlamydia trachomatis. For COVID-19 severity, only antibody titers loaded for finals models that more modestly predicted severe disease (AUC: 0.803; specificity=61.1%, sensitivity=85.7%). Nonetheless, this report shows that trait-like baseline data from 10-14 years ago can better characterize who is most at risk for COVID-19 and if they are likely to be hospitalized with a presumptively severe infection. In addition, our results suggest that past infection history and antibody response may be an invaluable, novel predictor of host immunity to COVID-19 that warrants further study.

## Methods

### Study design and participants

This retrospective study involved the UK Biobank cohort^12^. UK Biobank consists of approximately 500,000 people now aged 50 to 84 years (mean age=69.4 years). Baseline data was collected in 2006-2010 at 22 centers across the United Kingdom^13,14^. Summary data are listed in **Table 1**. This research involved deidentified epidemiological data. All UK Biobank participants gave written, informed consent. Ethics approval for the UK Biobank study was obtained from the National Health Service Health Research Authority North West - Haydock Research Ethics Committee (16/NW/0274). All analyses were conducted in line with UK Biobank requirements.

**Table 1.**
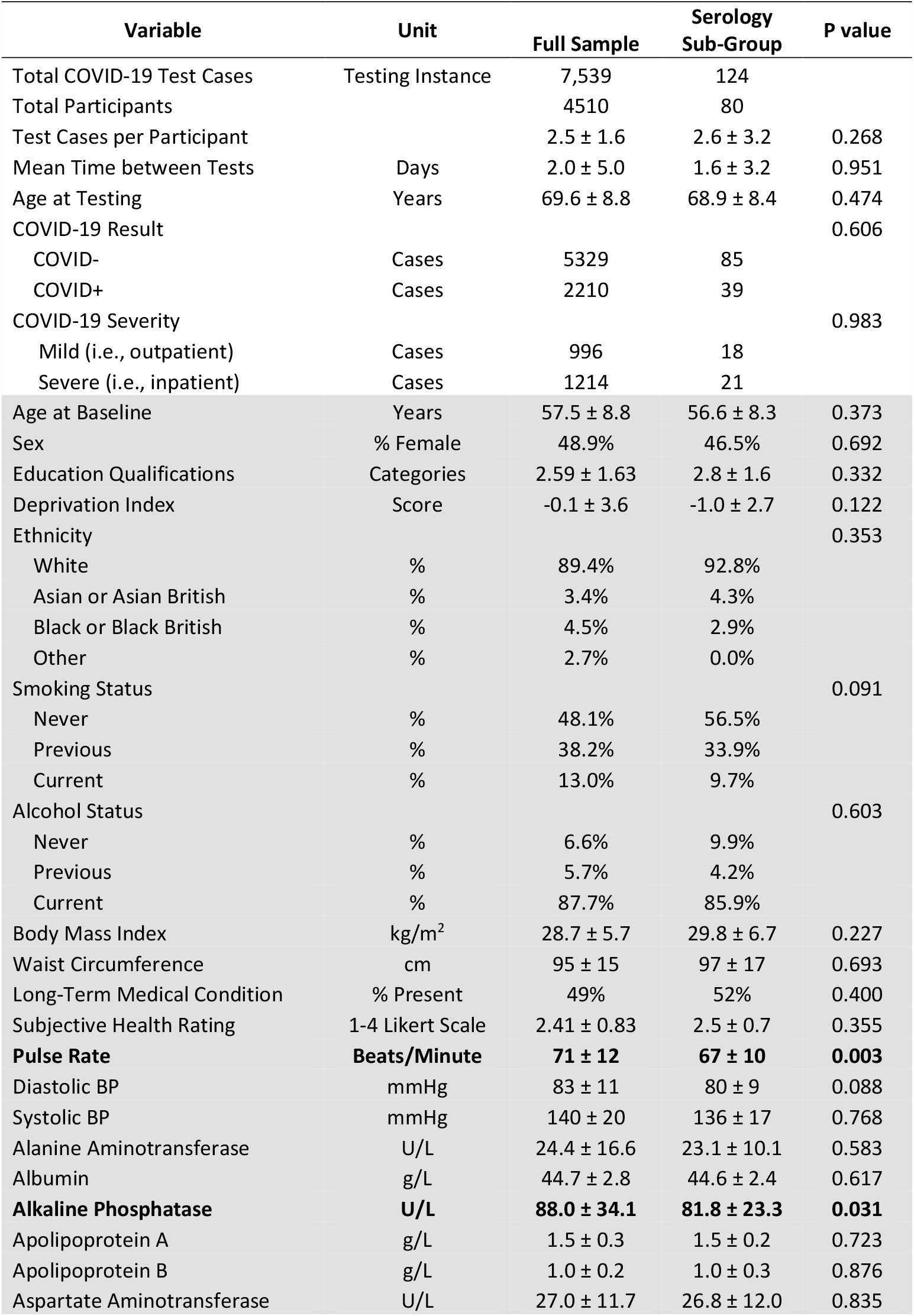
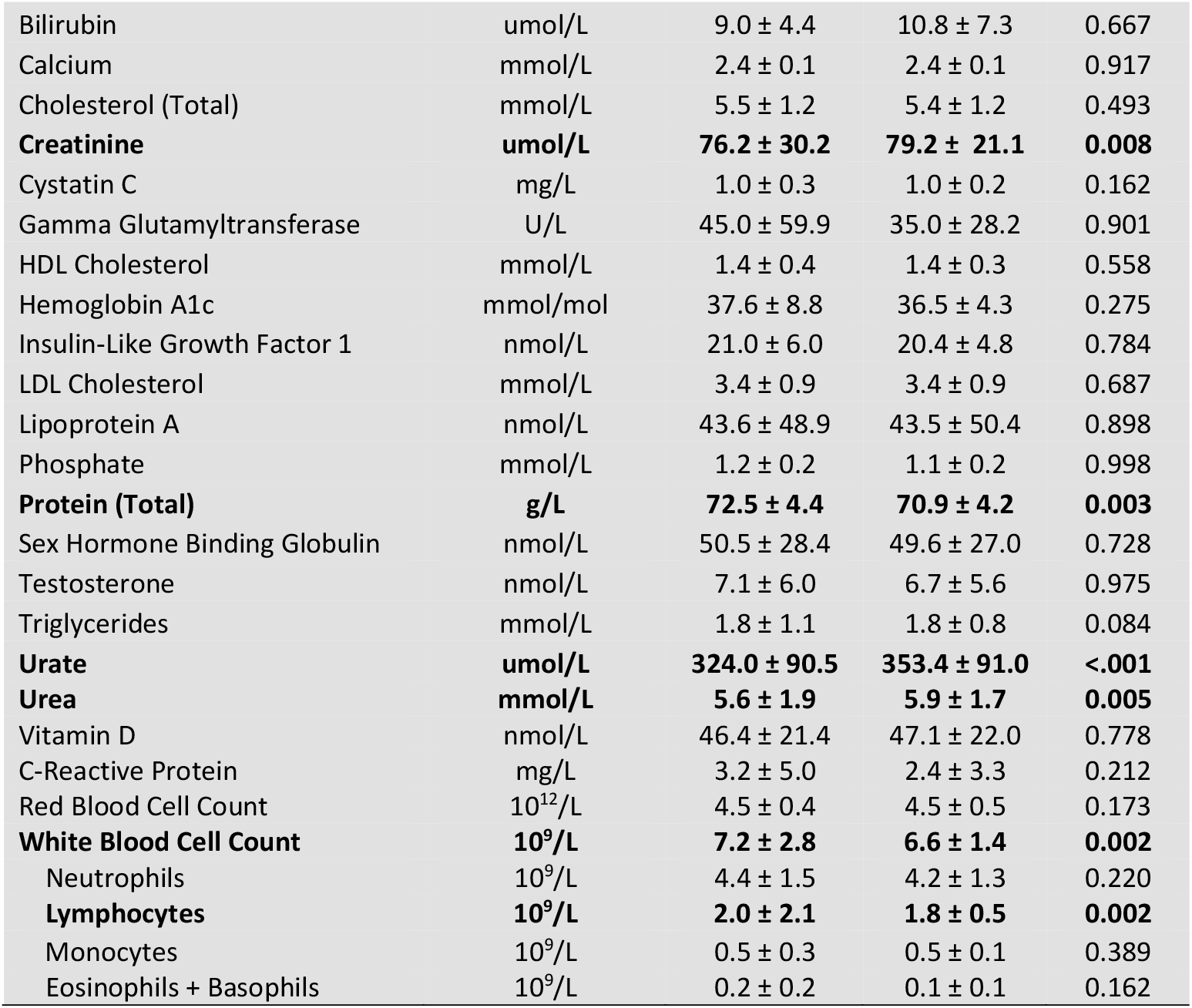
Baseline Demographics and Data Characteristics. Blood pressure (BP); high-density lipoprotein (HDL); low-density lipoprotein (LDL). A summary and comparison of data among either all participant test cases or a sub-group of test cases that also had non COVID-19 serology. Contemporary COVID-19 testing data has no shading. All retrospective baseline data has “gray” shading. Values are in Mean ± SD, percentages, or frequency. P values less than .05 were considered significant and applicable predictors and indices are bolded.

The following categories of predictors were downloaded: 1) demographics; 2) health behaviors and long-term disability or illness status; 3) anthropometric and bioimpedance measures of fat, muscle, or water content; 4) pulse and blood pressure; 5) a serum panel of thirty biochemistry markers commonly collected in a clinic or hospital setting; and 6) a complete blood count with a manual differential.

### Demographics

These factors included participant age in years at baseline, sex, education qualifications, ethnicity, and Townsend Deprivation Index. Sex was coded as 0 for female and 1 for male. For education, higher scores roughly correspond to progressively more skilled trade/vocational or academic training. Ethnicity was coded as UK citizens who identified as White, Black/Black British, or Asian/Asian British. The Townsend index^15^ is a standardized score indicating relative degree of deprivation or poverty based on permanent address.

### Health Behaviors and Conditions

This category consisted of self-reported alcohol status, smoking status, a subjective health rating on a 1-4 Likert scale (“Excellent” to “Poor”), and whether the participant had a self-described long-term medical condition. As noted in **Table 1**, 48.4% of participants indicated having such an ailment. We independently confirmed self-reported data with ICD-10 codes while at hospital. These conditions included all-cause dementia and other neurological disorders, various cancers, major depressive disorder, cardiovascular or cerebrovascular diseases and events, cardiometabolic diseases (e.g., type 2 diabetes), renal and pulmonary diseases, and other so-called pre-existing conditions.

### Vital Signs

The first automated reading of pulse, diastolic and systolic blood pressure at the baseline visit were used.

### Body Morphometrics and Compartment Mass

Anthropometric measures of adiposity (Body Mass Index, waist circumference) were derived as described^16^. Data also included bioelectrical impedance metrics that estimate central body cavity (i.e., trunk) and whole body fat mass, fat-free muscle mass, or water content^17^.

### Blood Biochemistry and Immunology

Serum biomarkers were assayed from baseline samples as described^18^. Briefly, using immunoassay or clinical chemistry devices, spectrophotometry was used to initially quantify values for 34 biochemistry analytes. UK Biobank deemed 30 of these markers to be suitably robust. We rejected a further 4 markers due data missingness >70% (estradiol, rheumatoid factor), or because there was strong overlap with multicollinear variables that had more stable distributions or trait-like qualities (glucose rejected vs. glycated hemoglobin/hba1c; direct bilirubin rejected vs. total bilirubin). A complete blood count with a manual differential was separately processed for red and white blood cell counts, as well as white cell sub-types.

### Serology Measures for Non COVID-19 Infectious Diseases

As described (http://biobank.ctsu.ox.ac.uk/crystal/crystal/docs/infdisease.pdf), among 9,695 randomized UK Biobank participants selected from the full 500,000 participant cohort, baseline serum was thawed and pathogen-specific assays run in parallel using flow cytometry on a Luminex bead platform^19^.

Here, the goal of the multiplex serology panel was to measure multiple antibodies against several antigens for different pathogens, reducing noise and estimating the prevalence of prior infection and seroconversion in at least UK Biobank. All measures were initially confirmed in serum samples using gold-standard assays with median sensitivity and specificity of 97.0% and 93.7%, respectively. Antibody load for each pathogen-specific antigen was quantified using median fluorescence intensity (MFI). Because seropositivity is difficult to assess for several pathogens, we did not use pathogen prevalence as a predictor in models.

**Table 2** shows the selected pathogens, their respective antigens, estimated prevalence of each pathogen based roughly on antibody titers, and assay values. This array ranges from delta-type retroviruses like human T-cell lymphotropic virus 1 that are rare (<1%) to human herpesviruses 6 and 7 that have an estimated prevalence of more than 90%.

**Table 2.**
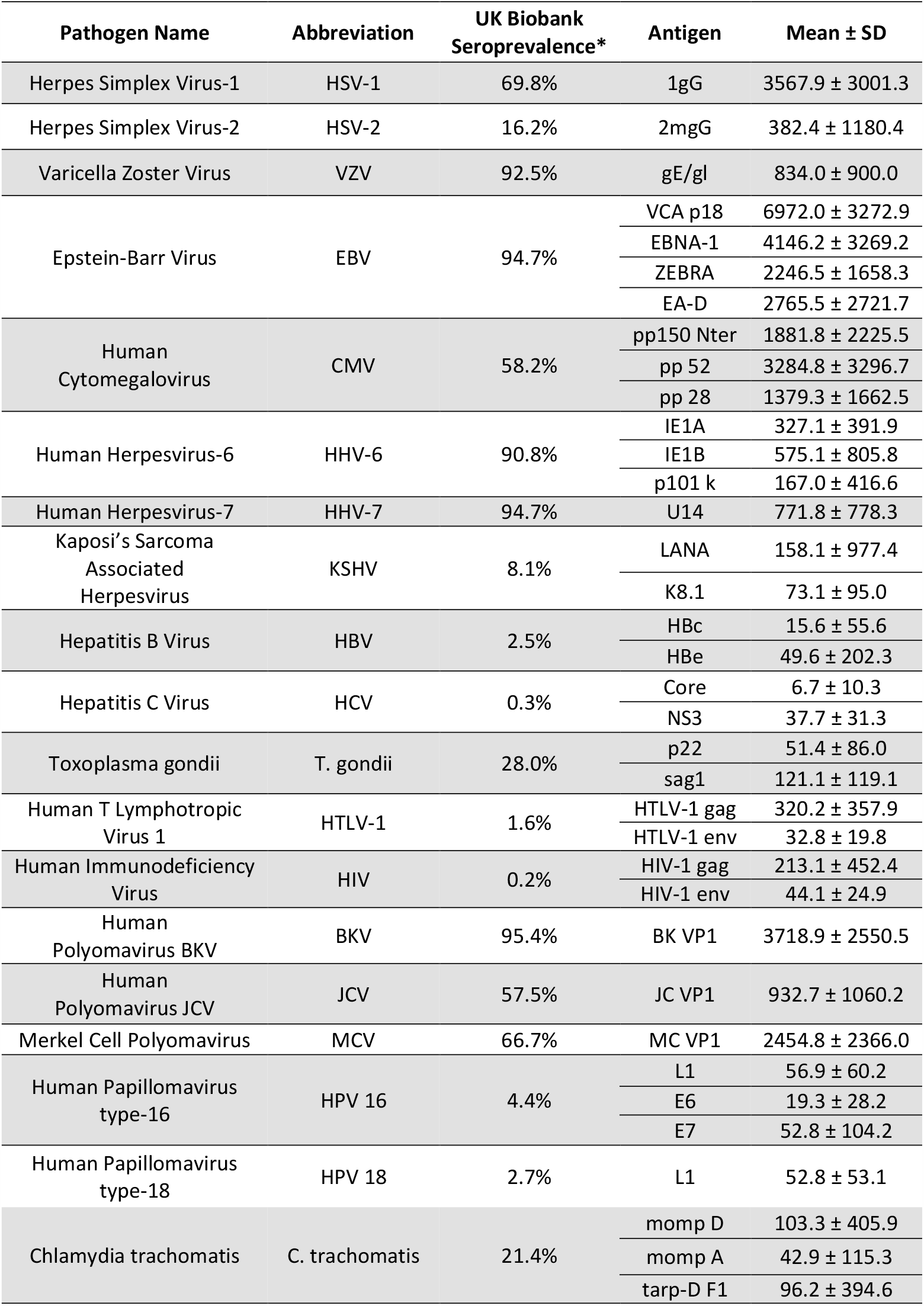
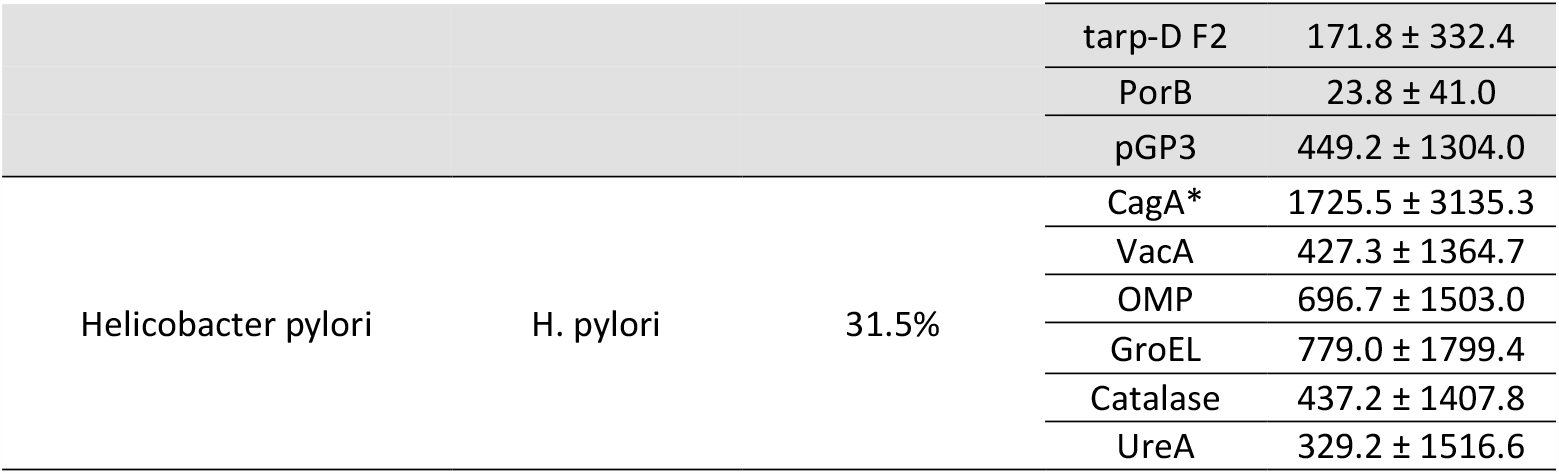
Baseline characteristics of infectious disease serology from 2006-2010. Antibody levels are specific to each antigen and expressed in Median Fluorescence Intensity (MFI) units. Seroprevalence of at least the main UK Biobank cohort was estimated on samples from 9,695 randomized participants, as described in white papers (see Methods). The “gray” and “white” shading are used to distinguish between pathogens and their respective antigens. *CagA levels are based on roughly half of the original sample due to a technical lab error.

### COVID-19 Testing

Our study was based on COVID PCR test data available from March 16^th^ to May 19^th^ 2020. Specifically, we used the May 26^th^, 2020 tranche of COVID-19 polymerase chain reaction (PCR) data from Public Health England. There were 4,510 unique participants that had 7,539 individual tests administered, hereafter called test cases. For modeling COVID-19 infection data, each test case was coded as ‘0’ and ‘1’, respectively representing a negative or positive PCR test. For modeling COVID-19 disease severity, each test case was coded as ‘0’ and ‘1’, which represented out-patient testing (i.e., presumptively mild case) or hospital in-patient testing with clinical signs of infection (i.e., presumptively severe case).

### Statistical Analyses

For a more technical description of the specific machine learning algorithm used to classify test cases, see **Supplemental Text 1**. SPSS 27 was used for all analyses and Alpha set at .05. Preliminary findings suggested that baseline serology data performed well in classifier models, despite a limited number of participants with serology. To determine if this serology sub-group was noticeably different from the full sample, Mann-Whitney U and Kruskal-Wallis tests were done (Alpha=.05). Hereafter, separate sets of classification analyses were performed for: 1) the full cohort; and 2) the sub-group of participants that had serology data. In other words, due to the imbalance of sample sizes and by definition the absence or presence of serology data, classifier performance in the serology sub-group was never statistically compared to the full cohort.

Next, linear discriminant analysis (LDA) was used to create predictive models that discriminated between negative vs. positive COVID-19 diagnosis or mild vs. severe disease status. LDA is a regression-like classification technique that finds the best linear combination of predictors that can maximally distinguish between groups of interest. To determine how useful a given predictor or related group of predictors (e.g., demographics) were for classification, simple forced entry models were first done. Subsequently, to derive “best fit,” robust models of the data, stepwise entry (Wilks’ Lambda, F value entry=3.84) was used to exclude predictors that did not significantly account for unique variance in the classification model. This data reduction step is critical because LDA can lead to model overfitting when there are too many predictors relative to observations^20,21^, which are COVID-19 test cases for our purposes. Finally, because there were multiple test cases that could occur for the same participant, this would violate the assumption of independence. To guard against this problem, we used Mundry and Sommer’s permutation LDA approach. Specifically, for each LDA model, permutation testing (1,000 iterations, P<.05) was done by randomizing participants across groupings of test cases to confirm robustness of the original model^22^.

LDA model overfitting can also occur when there is a sample size imbalance. Because there were many more negative vs. positive COVID-19 test cases in the full sample (5,329 vs. 2210), the negative test group was undersampled. Specifically, a random number generator was used to discard 2,500 negative test cases at random, such that the proportion of negative to positive tests was now 55% to 45% instead of 70.6% to 29.4%. Results without undersampling were similar (data not shown). No such imbalance was seen for COVID-19 severity in the full sample or for the serology sub-group. A typical holdout method of 70% and 30% was used for classifier training and then testing^23^. Finally, a two-layer non-parametric approach was used to determine model significance and estimated fit of one or more predictors. First, bootstrapping^24^ (95% Confidence Interval, 1000 iterations) was done to derive estimates robust against any violations of parametric assumptions. Next, ‘leave-one-out’ cross-validation^20^ was done with bootstrap-derived estimates to ensure that models themselves were robust. Collectively, the stepwise LDA models ensured that estimation bias of coefficients would be low because most predictors are “thrown out” before models are generated using the remaining predictors.

For each LDA classification model, outcome threshold metrics included: specificity (i.e., true negatives correctly identified), sensitivity (i.e., true positives correctly identified), and the geometric mean (i.e., how well the model predicted both true negatives and positives). The area under the curve (AUC) with a 95% confidence interval (CI) was reported to show how well a given model could distinguish between a COVID-19 negative or positive test result, and separately for COVID-19+ test cases if the disease was mild or severe. Receiver operating characteristic (ROC) curves plotted sensitivity against 1-specificity to better visualize results for sets of predictors and a final stepwise model. For stepwise models, the Wilks’ Lambda statistic and standardized coefficients are reported to see how important a given predictor was for the model. A lower Wilks’ Lambda corresponds to a stronger influence on the canonical classifier.

## Results

As shown in **Table 1**, 7,539 total test cases for COVID-19 were conducted among 4,510 UK Biobank participants (69.6 ± 8.8 years) between March 16^th^ to May 19^th^ 2020, either in outpatient or inpatient settings. There were 5,329 negative cases and 2,210 positive cases. Of the positive cases, there were 996 mild and 1,214 presumptively severe disease outcomes. Baseline data from 10-14 years ago (Mean = 11.22 years) was available for demographic, laboratory, biochemistry, and clinical indices. Similar data from 2020 was not available. A central theme of this report is examining prediction models for the so-called full sample, but also an entirely separate set of models for a sub-group of test cases with serology data (**Table 2**). **Table 1** indicates that the full cohort and serology sub-groups largely did not differ on most measures. A few significant differences were clinically unremarkable for the serology sub-cohort and well within the range of normal values, including lower pulse rate, several markers reflecting better kidney function, and lower total white blood cell count due to fewer lymphocytes.

Next, each baseline variable was used to predict COVID-19 infection for a given test case. For context, model performance was judged by: 1) the AUC as a measure of probability, where 0.5 is at-chance prediction and 1.0 is perfect prediction; and 2) the geometric mean or g-mean as a threshold metric, with a higher percentage corresponding to greater likelihood of correctly identifying both true positives and true negatives. Among all participants (**Supplementary Table 1**), as expected, model fit was poor for individual predictors that loaded significantly (mean AUC=0.532; AUC range=0.517-0.551). For example, known risk factors included larger body composition indices (AUC=0.526-0.548; g-mean=16.2%-29.6%), older age (AUC=0.522; g-mean=38.8%), and markers of dysmetabolism like higher hba1c % (AUC=0.537; g-mean=13.3%) and high diastolic blood pressure (AUC=0.519; g-mean=17.9%).

For the serology sub-group (**Supplementary Table 2**), several established risk factors that loaded had better overall fit (mean AUC=0.656, AUC range=0.601-0.731). Like the full sample, examples included larger body mass like fat-free mass (AUC=0.687; g-mean=65.0%), hba1c % (AUC=0.638; g-mean=52.8%), and diastolic blood pressure (AUC=0.633; g-mean=55.2%). Some unexpected factors included total protein (AUC=0.662; 65.8%) and testosterone (AUC=0.731; g-mean=55.8%). We then tested if antibody titers to antigens of 20 rare to common infectious pathogens could predict host immunity in 2020 to COVID-19. As shown in **Supplementary Table 3**, antibody titers to 15 antigens across 12 pathogens each performed as well on average as other non-serology predictors (mean AUC=0.653, AUC range=0.612-0.710). Specificity and sensitivity were notable for antibody levels to the pp150 Nter antigen to human cytomegalovirus (g-mean=61.0%) and U14 to Human Herpes Virus-7 (g-mean=66.1%), given their prevalence in the sample.

Next, sets of similar predictors were used to gauge how well they collectively predicted COVID-19 infection, as listed in **Table 3** and shown using ROC curves in **Figure 1**. A stepwise model was also used to create a classifier that only included predictors where each provided unique predictive utility, and to minimize likelihood of overfitting models. For the full sample (**Table 3**, top row), sets of predictors including the stepwise model were able to identify COVID-19 negative and positive test cases up to 96.1% and 23.8% of the time respectively. **Supplementary Table 4** (top row) illustrates that the stepwise model included triglycerides, body mass, age, ethnicity, and other known risk factors for COVID-19. Importantly, for the serology sub-group (**Table 3**, bottom row), forced entry models showed worse performance compared to the same models among the full sample, except for the biochemistry set. This suggests that small sample size for the serology sub-group did not lead to model overfitting. While the forced entry serology model itself is likely overfitted, the stepwise model loaded 15 predictors and performed well (g-mean=0.920). As shown in **Supplementary Table 4** (bottom row), predictors that loaded in the stepwise model included antibody titers for antigens of several common pathogens (e.g., human cytomegalovirus, C. trachomatis), lipid markers, age, white and red cell counts, and testosterone. Due to potential concerns with model overfitting, the stepwise model was re-run with only predictors that had individually loaded significantly in forced entry models (**Supplementary Tables 2 and 3**). This stepwise model had 10 variables and achieved a g-mean of 85.4%, suggesting that stepwise models were not overfitted.

**Table 3.** Sets of predictors used to predict classification of COVID-19 test cases as negative or positive. Area Under the Curve (AUC); Confidence Interval (CI); Geometric Mean (G-Mean). Non-parametric bootstrapping (1000 iterations, 95% CI) was used for robust estimation. P values less than .05 were considered significant. “Blue” and “white” shading are used to distinguish between predictors that loaded for a given model. *Due to several variables representing the same construct (i.e., being multicollinear), body composition consisted of: whole-body water mass; whole-body fat mass; whole-body non-fat mass (i.e., muscle, bone).

|  | Sets of Predictors | Number of Predictors | Classifier Method | P value | AUC (95% CI) | Specificity | Sensitivity | G-Mean |
| --- | --- | --- | --- | --- | --- | --- | --- | --- |
| Full Sample | Basic Demographics | 5 | Enter | <.001 | 0.539 (0.557-0.520) | 92.4% | 12.4% | 0.338 |
|  | Body Composition* | 3 | Enter | <.001 | 0.536 (0.552-0.520) | 92.7% | 8.5% | 0.281 |
|  | Health Behaviors/Conditions | 4 | Enter | .011 | 0.527 (0.511-0.544) | 93.5% | 8.2% | 0.277 |
|  | Vitals | 3 | Enter | .004 | 0.526 (0.510-0.542) | 96.1% | 4.8% | 0.215 |
|  | Biochemistry | 26 | Enter | .001 | 0.575 (0.555-0.596) | 81.2% | 22.9% | 0.431 |
|  | Immunology | 8 | Enter | <.001 | 0.533 (0.517-0.549) | 94.4% | 7.7% | 0.270 |
|  | Stepwise Model | 8 | Stepwise | <.001 | 0.549 (0.419-0.680) | 83.0% | 23.8% | 0.444 |
| Serology Sub-Group | Basic Demographics | 5 | Enter | 0.008 | 0.713 (0.620-0.805) | 81.2% | 10.3% | 0.289 |
|  | Body Composition* | 3 | Enter | 0.013 | 0.660 (0.558-0.763) | 96.5% | 5.1% | 0.222 |
|  | Health Behaviors/Conditions | 4 | Enter | 0.006 | 0.694 (0.599-0.789) | 98.8% | 0% | 0 |
|  | Vitals | 3 | Enter | 0.261 | 0.593 (0.485-0.702) | 98.8% | 0% | 0 |
|  | Biochemistry | 26 | Enter | <.001 | 0.799 (0.698-0.900) | 80.0% | 56.4% | 0.672 |
|  | Immunology | 8 | Enter | 0.393 | 0.639 (0.540-0.737) | 94.1% | 5.1% | 0.219 |
|  | Serology | 44 | Enter | <.001 | 0.976 (0.952-1.000) | 80.0% | 76.9% | 0.784 |
|  | Stepwise Model | 15 | Stepwise | <.001 | 0.969 (0.934-1.000) | 91.8% | 92.3% | 0.920 |

**Figure 1.**
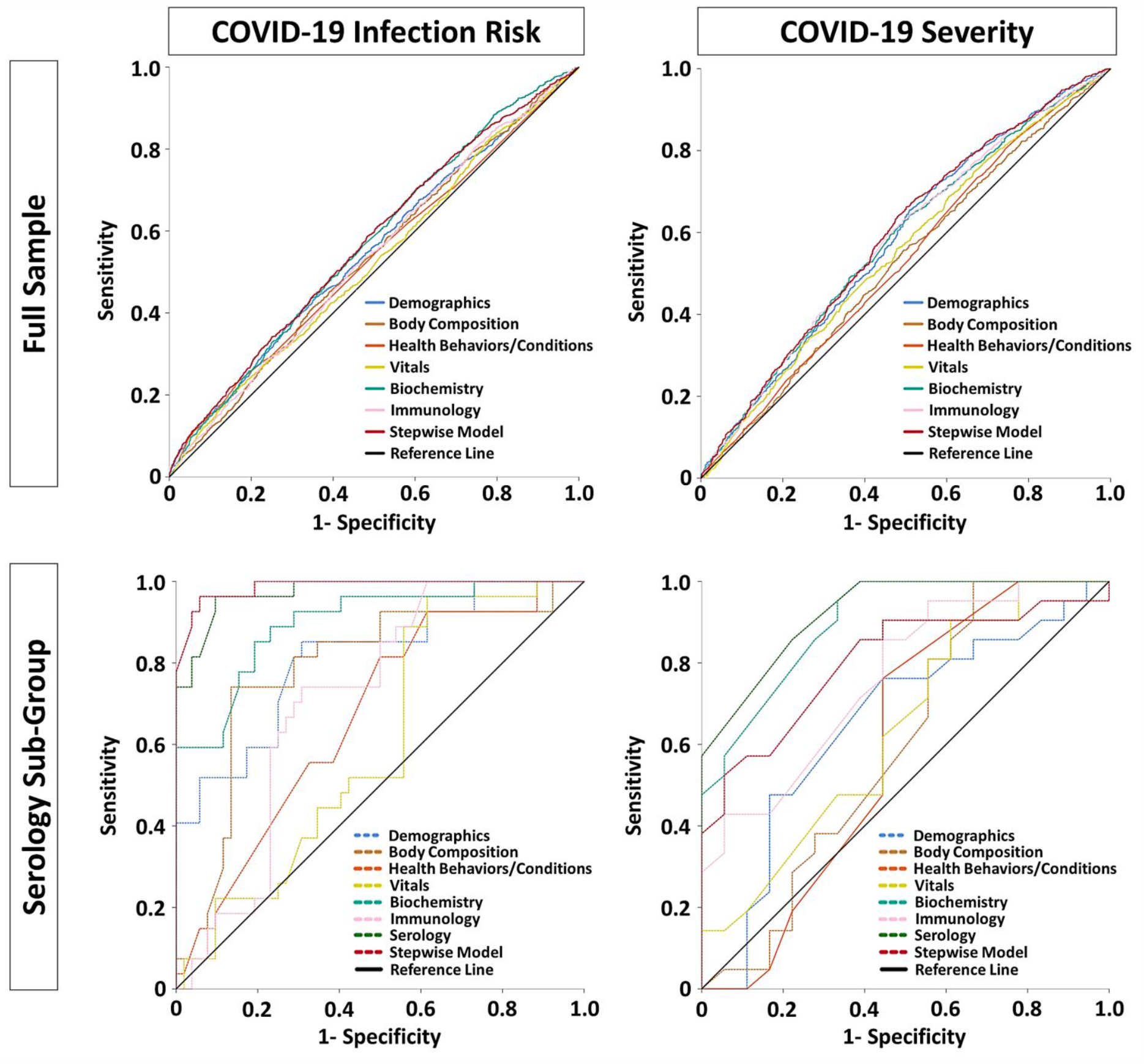
ROC curves and model fit for COVID-19 infection risk and infection severity. Receiver Operating Characteristics (ROC) curves illustrating the relative classifier performance of various sets of predictors. Outcomes of interest were COVID-19 infection risk and whether an infection was mild or severe. Two separate sets of analyses were done for the full tested sample and a sub-group of participants with serology data. Test statistics for predictors are provided in Tables 3 and 4.

Separately, another set of analyses determined how each baseline predictor could predict which of the 2,210 positive COVID-19 cases had a mild or severe disease course. For context, 45% and 55% of test cases were mild or severe respectively. Among all positive test cases (**Supplementary Table 5**), significant predictors showed a trade-off between better sensitivity or specificity and were only modestly useful (AUC mean and range=0.536, 0.524-0.572). Similarly, for the serology sub-group, **Supplementary Table 6** shows that only alanine aminotransferase and neutrophil count significantly predicted disease severity. For serology data itself, **Supplementary Table 7** indicates that the only significant predictors were U14 antigen to human herpesvirus 7 (AUC=0.729; g-mean=0.600) and JC VP1 antigen to human JC polyomavirus (AUC=0.671; g-mean=0.591). **Table 4** shows the relative predictive value of groups of predictors for whether a COVID-19 infection would be severe. **Figure 1** shows the ROC curves for model fit. **Supplementary Table 8** illustrates that the stepwise model included only alanine aminotransferase, age in years, and monocyte count. For the serology sub-group, despite strong concerns about model overfitting, the AUC and g-mean were similarly modest compared to the full sample, except for the stepwise model that performed noticeably better (AUC=0.803; g-mean=0.724). As shown in **Supplementary Table 8**, this model had only 2 predictors: antibody response to two antigens for two diseases (HTLV-1 gag for HTLV-1 and JC VP1 for Human Polyomavirus JCV).

**Table 4.** Sets of predictors used to predict classification of COVID-19 positive cases as mild or severe. Area Under the Curve (AUC); Confidence Interval (CI); Geometric Mean (G-Mean). Non-parametric bootstrapping (1000 iterations, 95% CI) was used for robust estimation. P values less than .05 were considered significant. “Orange” and “white” shading are used to distinguish between predictors that loaded for a given model. *Due to several variables representing the same construct (i.e., being multicollinear), body composition consisted of: whole-body water mass; whole-body fat mass; whole-body non-fat mass (i.e., muscle, bone). ^= Due to the full serology panel of 44 antibody titers exceeding degrees of freedom, titers for 6 antigens were excluded for pathogens with the lowest estimated prevalence in the cohort (HIV, HCV, HTLV-1).

|  | Sets of Predictors | Number of Predictors | Classifier Method | P value | AUC (95% CI) | Specificity | Sensitivity | G-Mean |
| --- | --- | --- | --- | --- | --- | --- | --- | --- |
| Full Sample | Basic Demographics | 5 | Enter | <.001 | 0.581 (0.557-0.605) | 36.9% | 74.6% | 0.525 |
|  | Body Composition* | 3 | Enter | 0.025 | 0.528 (0.504-0.552) | 0.9% | 99.5% | 0.095 |
|  | Health Behaviors/Conditions | 4 | Enter | 0.011 | 0.531 (0.507-0.556) | 0.0% | 96.3% | 0 |
|  | Vitals | 3 | Enter | <.001 | 0.554 (0.530-0.578) | 7.0% | 94.6% | 0.257 |
|  | Biochemistry | 26 | Enter | <.001 | 0.579 (0.555-0.602) | 22.1% | 81.5% | 0.424 |
|  | Immunology | 8 | Enter | <.001 | 0.581 (0.557-0.605) | 5.5% | 95.8% | 0.230 |
|  | Stepwise Model | 3 | Stepwise | <.001 | 0.592 (0.568-0.615) | 36.4% | 76.1% | 0.526 |
| Serology Sub-Group | Basic Demographics | 5 | Enter | 0.964 | 0.652 (0.472-0.832) | 22.2% | 47.6% | 0.325 |
|  | Body Composition* | 3 | Enter | 0.665 | 0.597 (0.407-0.786) | 33.3% | 81.0% | 0.519 |
|  | Health Behaviors/Conditions | 4 | Enter | 0.994 | 0.598 (0.404-0.792) | 35.3% | 14.3% | 0.225 |
|  | Vitals | 3 | Enter | 0.448 | 0.636 (0.459-0.814) | 44.4% | 66.7% | 0.544 |
|  | Biochemistry | 26 | Enter | <.001 | 0.901 (0.808-0.993) | 33.3% | 71.4% | 0.488 |
|  | Immunology | 8 | Enter | <.001 | 0.763 (0.615-0.911) | 44.4% | 71.4% | 0.563 |
|  | Serology^ | 36 | Enter | <.001 | 0.925 (0.847-1.000) | 38.9% | 81.0% | 0.561 |
|  | Stepwise Model | 2 | Stepwise | <.001 | 0.803 (0.663-0.943) | 61.1% | 85.7% | 0.724 |

## Discussion

The objectives of this study were to determine if baseline data from 2006-2010 could predict which older adults would develop COVID-19 in 2020, and if an infection was mild or presumptively severe due to being at hospital. In summary, using a permutation-based LDA approach, we developed separate risk profiles that did well at predicting test cases that were negative or positive (stepwise g-mean=92%), and to some degree among positive test cases whether the infection was mild or severe (stepwise g-mean=72.4%). Such profiles would require retrospective, routine self-report, blood test panels typically collected during annual medical wellness visits, and serology information for several antigens. As proof-of-principle that these profiles are sensible, we confirmed as others have noted that non-white ethnicity, low socioeconomic status, larger body mass, and alcohol use can increase infectious risk^5^.

Our most novel finding was that antibody titers to past infections were strong predictors of COVID-19 infection and severity, both as a group and especially in concert with established risk factors. This “virome” may consist of beneficial and detrimental pathogens, or fine-grained efficacy of the immune system to clear certain pathogens, that change how the immune system responds to a persistent viral challenge like COVID-19^10^. For example, antibodies to human cytomegalovirus antigens were the strongest predictors of infection risk in our stepwise model. Older adults with prior infection show exhaustion of the naïve T cell pool and fewer memory versus effector cells^25^. This may explain why monocyte count was one of the few variables to predict COVID-19 severity among all test cases in this study, as innate immunity must compensate for deficits in acquired immune function. For COVID-19 severity, antibody titers to the HTLV1 virus and human JC polyomavirus were the only predictors that loaded significantly in our stepwise model. While HTLV1 is rare, 57.5% of at least the UK Biobank sample have antibody levels that suggest prior infection with the human JC polyomavirus. This virus can induce hemagglutination in type O blood cells^26^, which may in some way influence why this blood type is protective against COVID-19 infection.

For other immunologic factors, as expected, mobilization of innate immunity was relevant to infection risk and severity. In particular, granulocytes (e.g., neutrophils, monocytes) loaded significantly in stepwise models for COVID-19 infection and severity, but not cytokines such as C-Reactive Protein. C-Reactive Protein has been cited as a strong risk factor for COVID-19^27^. However, this marker merely reflects signaling of the acute phase response due to systemic infection, and changes to granulocytes in circulation already reflect this response. Although lymphopenia and suppression of humoral immunity have been noted in COVID-19, lymphocyte cell count did not load in final stepwise models.

We also confirmed and extended the importance of age, lipids, vascular health, and socioeconomic status, but while body mass was important it was not adiposity per se. Among mostly elderly adults in our UK Biobank sample, age was one of the few factors to impact both infection and severity risk. Perhaps in concert, lipoprotein metabolism changes with aging can induce hyperlipidemia, which is a risk factor for cardiovascular disease and may increase COVID-19 infection risk^28^. The lack of association with anthropometric or bioimpedance-derived fat mass was unexpected, whereas fat-free mass such as muscle and bone did load as a factor. We speculate that more bone mineral density and somatic muscle would reflect less cardiometabolic impairment and systemic inflammation, but mechanisms are unclear. Finally, levels of testosterone weakly loaded as a predictive factor for who would later develop COVID-19. Sex differences favoring COVID-19 infection in men are clear, bout andropause induces less testosterone production, which normally downregulates inflammation, and could increase COVID-19 susceptibility^29^.

Several major limitations should be noted. The number of UK Biobank participants with COVID-19 and serology data is low, particularly for positive test cases. This could consequently lead to model overfitting or misestimation. Several steps were taken to guard against this problem, including feature reduction through LDA, bootstrapped parameter estimation to guard against parametric assumption violations, and several cross-validation steps to maximize robustness. We also rigorously tested each predictor or set of predictors in the main sample and serology sub-group, where we found that model fit was not overly biased in general despite sample size differences. Nonetheless, we recognize future work must use much larger sample sizes to verify the usefulness of serology data. Another limitation was that using test case data nested within a participant violates the assumption of independence, which can lead to gross misestimation. While we ameliorated this issue using permutation testing, other latent concerns with the data like type 2 error may be present. We also chose to use LDA over other machine learning algorithms, where LDA tends to provide more conservative estimates. This was intentional, because it is still largely unknown how risk factors alone or additively reflect overall risk for COVID-19 infection and disease severity. Finally, we only looked at the so called main effects of all predictors instead of complex interactions, such as darker skin, vitamin D levels, and COVID-19 infection risk. Such interactions were beyond the scope of this report, but may be promising avenues to explore in future studies.

## Conclusions

In summary, this study systematically used retrospective data in a large community cohort to predict who would develop COVID-19 and if the disease course was presumptively severe. Despite baseline data having been collected 10-14 years ago, we achieved excellent to encouraging results by combining several sets of established and novel risk factors together. It is especially interesting that serological data performed as well as or better than other data types. Future work should leverage markers of host immunity to inform what may happen when the host is challenged by COVID-19.

## Data Availability

All data was downloaded from the UK Biobank, a data resource open to all bona fide researchers.

## Additional Information

### Author Contribution Statement

AAW performed literature searches, created all figures and tables, formed the study design, analyzed and interpreted the data, and wrote the manuscript. SAW performed literature searches, helped form the study design, interpreted the data, and edited the manuscript. QW and CP managed most of data collection, interpreted the data, and edited the manuscript. BSK, SL, BL, TL, and AP managed part of data collection and interpreted the data. NB and TW originally acquired part of the data (serology) and interpreted the data.

### Data Availability

The data that support the findings of this study are available from the UK Biobank but restrictions apply to the availability of these data, which were used under license for the current study, and so are not publicly available. Data are however available from the authors upon reasonable request and with permission of UK Biobank. Permission is acquired through an application for data access to UK Biobank, https://www.ukbiobank.ac.uk/register-apply/.

### Ethics Declarations

Ethics approval for the UK Biobank study was obtained from the National Health Service Health Research Authority North West - Haydock Research Ethics Committee (16/NW/0274). All analyses were conducted in line with UK Biobank requirements.

## Supplemental Text 1

To reiterate, the main objectives of this report are to use Linear Discriminant Analysis (LDA) for probabilistic determination via naïve Bayesian classification of: 1) a two-class grouping defined as a positive vs. negative COVID-19 test case; and 2) a two-class grouping nested within positive COVID-19 tests, defined as a test case occurring in a hospital vs. non-hospital setting. For positive tests, these settings are considered as proxies for mild vs. severe COVID-19 disease status. Because two or more test cases could be nested within a given participant, this would normally violate independence and potentially invalidate results. While one could use a single random test case per participant, this reduces sample size and does not represent real world data and within-subject variability (e.g., changes in COVID-19 status or infection severity). Thus, for estimation robust to non-independence, we used Mundry and Sommer’s permuted LDA approach^22^. The unit of randomization across participants was a grouping of all test cases originally nested in a given participant. The null hypothesis was that a given LDA model with randomized data would not perform better than the original non-randomized data in 95% of permutations. As recommended, 1,000 permutations were run for each model using macros provided by the authors and Python scripting for automation. To ensure that models were stable and generalizable, we used a typical “holdout” method of 70% and 30% respectively for the training and test samples^23^.

As discussed in the main text, forced entry models were first conducted for each predictor, and then among sets of similar predictors (e.g., demographics, vital signs). It was known from the outset that model overfitting would likely occur when a predictor set had dozens of features (e.g., biochemistry). This procedure was done for two reasons: 1) to show clinicians, researchers, and policymakers how a set of common features would discriminate COVID-19 groups in ideal circumstances, where in some cases model overfit is frankly likely; and 2) to contrast such models with the comparable or superior performance of stepwise models that had substantial feature reduction and enough n > p that model overfit was guarded against.

We now explain why LDA was used. Foremost, prediction models derived using LDA are straightforward to interpret by a general audience, which is appropriate for the journal in question. For example, the Wilks’ Lambda statistic allows a clear interpretation for how well a given predictor distinguishes between classes and its directionality (e.g., higher age in years predicts increased likelihood of positive COVID-19 classification).

Equally important, LDA creates models that maximally separate classes of interest, where a new observation’s data can be used to determine which class that observation would belong in. Since it is of central importance to have equally valid diagnostic assessment for who is and is not at risk for COVID-19 or if a positive would have a mild or severe infection, LDA is most appropriate. As a generative, supervised learning classification technique, LDA is also best used in complex datasets with high dimensionality composed of a few to hundreds of features per data category, where it can remove most redundant or dependent features that do not maximize model fit. Reducing the feature set size reduces the risk of model overfitting^20,21^. This procedure minimizes the High Dimensional, Low Sample Size (i.e., p>>N, or “small *n*, big *p*”) problem^20^ by reducing the likelihood of the within-class covariance matrix approaching singularity^30^ and leading to instability of parameter estimation.

LDA has several key assumptions that we wish to address. LDA is relatively robust to overfitting provided there is a relative lack of outliers, multivariate normality and lack of multicollinearity, and independence of data values between participants. To begin, UK Biobank removes extreme values during data quality control before posting datasets to their data showcase^12^. We further log-transformed all quantitative variables to normalize distributions and “bring in” outliers defined as data points >3SD from the mean. As described in the main text, we also removed biochemistry variables that were multicollinear (e.g., direct vs. total bilirubin). While some antigens of the same pathogen approached multicollinearity, removing them from feature selection led to identical results and thus they were kept in. Participant-level data was not dependent on data from other participants. To be clear, however, multiple observations were nested within a given participant and would violate independence. Because the permutation LDA testing randomizes which participants have a group of one to several COVID-19 test cases, however, these models are robust to non-independence.

While other machine learning techniques are also appropriate for classification, we discuss why they were not used. Regarding logistic regression, this technique is attractive because it has no distribution assumptions. However, it assumes observations are independent. This does not occur with COVID-19 testing, in which a participant will often have multiple test cases. Logistic regression also requires a large numbers of observations to provide reliable estimates. Finally, it does not produce robust models for well-separate types of classes. As there are very clear immunologic differences that determine if someone has or does not have COVID-19, and a clear demarcation between mild vs. severe symptom presentation, we believe logistic regression model estimations might be inflated and thus less accurate. Finally, despite their methodological differences, LDA and logistic regression may perform the same with real data^31^, where LDA may be more conservative and was one of our goals for this proof-of-principle study.

More complex algorithms vs. LDA were also not considered due to feature complexity, the need for transparent model estimates, and sample size. First, in the dataset there are many features present for biochemistry markers, antibody load to specific antigens, and to a lesser degree immune factors. Data reduction is therefore important to determine which features are most useful for COVID-19 data and should receive attention. By contrast, clustering methods are not suitable because the dimensionality space is too high and model fit is likely to be poor. For newer machine learning techniques, such as deep learning, it is often unclear what set of features are selected or their relative contribution when a given prediction is made. This is unacceptable for predicting COVID-19 infection or severity risk. For researchers, it is unknown how various risk factors converge to affect risk and this information is necessary to better understand underlying mechanisms. In population health or the clinic, certain features have prohibitive time or cost constraints (e.g., body compartment imaging; ordering one versus multiple antigen tests). More importantly, it is critical for clinicians, policy makers, or other stakeholders to point out which exact features led or would lead to a predicted outcome. Finally, deep learning, support vector machines (SVM), and similar approaches also require much larger sample sizes to train and adapt a classifier to produce robust estimates. By comparison, our dataset only had several thousand testing datapoints in the “full” sample and just over one-hundred in the sub-group that had serological data.

We recognize that LDA has several limitations and used non-parametric estimation to minimize these issues. To begin, using simulation data, LDA performs comparably to logistic regression when predictor distributions are normal or near normal, but has worse fit when there are clear normality violations^32^. While we log-transformed quantitative measures with appreciable skewness (>3SD), normality nonetheless remained a concern, particularly for the serology sub-group that had 124 observations. To reduce potential problems, bootstrapping^24^ was used (95% CI, 1000 iterations) to estimate model coefficients. This allows unbiased estimation of generalized absolute error, taking into account potential model overfit by substantially varying training and test sets from the selected sample. Nevertheless, with the serology sub-group, the small *n*, big *p* problem may still be a concern. Regularized LDA has been a popular choice to overcome this issue of within-class covariance singularity, where cross-validation presents a reasonable solution^33^. Due to computation problems in tandem with bootstrapping, we used a simple “leave-one-out” approach with bootstrap estimates.

**Supplementary Table 1.** Isolated effect of each non-serology predictor on COVID-19 risk among the full sample.

| Predictor | Classifier Method | P value | AUC (95% CI) | Specificity | Sensitivity | G-Mean |
| --- | --- | --- | --- | --- | --- | --- |
| **Basic Demographics** |  |  |  |  |  |  |
| Age | Enter | **0.002** | **0.522 (0.505-0.538)** | **86.9%** | **17.3%** | **0.388** |
| Sex | Enter | **0.033** | **0.517 (0.501-0.534)** | **100%** | **0%** | **0** |
| Ethnic Background | Enter | 0.080 | 0.514 (0.498-0.531) | 93.6% | 11.2% | 0.324 |
| Deprivation Index | Enter | **<.001** | **0.540 (0.524-0.556)** | **79.5%** | **22.5%** | **0.423** |
| Education | Enter | 0.626 | 0.505 (0.479-0.512) | 100% | 0% | 0 |
| **Body Composition** |  |  |  |  |  |  |
| Waist Circumference | Enter | **<.001** | **0.544 (0.528-0.561)** | **97.0%** | **2.7%** | **0.162** |
| Body Mass Index | Enter | **<.001** | **0.548 (0.531-0.564)** | **94.0%** | **6.4%** | **0.245** |
| Trunk Fat Mass | Enter | **<.001** | **0.533 (0.516-0.549)** | **97.4%** | **3.3%** | **0.179** |
| Whole Body Fat Mass | Enter | **0.002** | **0.526 (0.509-0.542)** | **91.1%** | **8.8%** | **0.283** |
| Whole Body Fat-Free Mass | Enter | **0.005** | **0.529 (0.513-0.545)** | **91.4%** | **9.6%** | **0.296** |
| Whole Body Water Mass | Enter | **0.005** | **0.529 (0.512-0.545)** | **91.5%** | **9.6%** | **0.296** |
| **Health Behaviors and Conditions** |  |  |  |  |  |  |
| Smoking Status | Enter | 0.167 | 0.511 (0.472-0.505) | 100% | 0% | 0 |
| Alcohol Status | Enter | **<.001** | **0.517 (0.501-0.533)** | **88.6%** | **14.6%** | **0.360** |
| Long-Term Medical Condition | Enter | 0.691 | 0.504 (0.488-0.520) | 100% | 0% | 0 |
| Health Rating | Enter | 0.354 | 0.503 (0.481-0.514) | 100% | 0% | 0 |
| **Vitals** |  |  |  |  |  |  |
| Pulse Rate | Enter | 0.954 | 0.503 (0.481-0.514) | 100% | 0% | 0 |
| Diastolic BP | Enter | **0.041** | **0.519 (0.503-0.536)** | **97.3%** | **3.3%** | **0.179** |
| Systolic BP | Enter | 0.952 | 0.501 (0.483-0.516) | 100% | 0% | 0 |
| **Biochemistry** |  |  |  |  |  |  |
| Alanine Aminotransferase | Enter | 0.302 | 0.512 (0.491-0.533) | 99.8% | 0% | 0 |
| Albumin | Enter | 0.154 | 0.504 (0.484-0.525) | 100% | 0% | 0 |
| Alkaline Phosphatase | Enter | 0.781 | 0.508 (0.487-0.528) | 100% | 0% | 0 |
| Apolipoprotein A | Enter | **<.001** | **0.536 (0.515-0.557)** | **96.4%** | **3.0%** | **0.170** |
| Apolipoprotein B | Enter | 0.192 | 0.514 (0.493-0.534) | 99.8% | 0.1% | 0.032 |
| Aspartate Aminotransferase | Enter | 0.284 | 0.518 (0.498-0.539) | 100% | 0% | 0 |
| Bilirubin (Total) | Enter | 0.377 | 0.501 (0.480-0.521) | 100% | 0.1% | 0.032 |
| Calcium | Enter | 0.104 | 0.517 (0.497-0.538) | 99.8% | 0.4% | 0.063 |
| Cholesterol (Total) | Enter | 0.833 | 0.517 (0.484-0.525) | 100% | 0% | 0 |
| Creatinine | Enter | 0.898 | 0.505 (0.515-0.556) | 100% | 0% | 0 |
| **Cystatin C** | **Enter** | **0.032** | **0.523 (0.502-0.543)** | **100%** | **0%** | **0** |
| Gamma Glutamyltransferase | Enter | 0.648 | 0.502 (0.481-0.522) | 100% | 0% | 0 |
| **HDL Cholesterol** | **Enter** | **<.001** | **0.536 (0.515-0.557)** | **95.2%** | **4.3%** | **0.202** |
| **Hemoglobin A1c** | **Enter** | **0.006** | **0.537 (0.516-0.558)** | **98.3%** | **1.8%** | **0.133** |
| Insulin-Like Growth Factor 1 | Enter | 0.316 | 0.504 (0.483-0.525) | 100% | 0% | 0 |
| LDL Cholesterol | Enter | 0.105 | 0.504 (0.484-0.525) | 100% | 0% | 0 |
| Lipoprotein A | Enter | 0.081 | 0.503 (0.482-0.524) | 100% | 0% | 0 |
| Phosphate | Enter | 0.173 | 0.515 (0.494-0.536) | 100% | 0% | 0 |
| Protein (Total) | Enter | 0.078 | 0.517 (0.497-0.538) | 100% | 0% | 0 |
| **Sex Hormone Binding Globulin** | **Enter** | **<.001** | **0.551 (0.531-0.572)** | **100%** | **0%** | **0** |
| Testosterone | Enter | 0.096 | 0.513 (0.492-0.533) | 100% | 0% | 0 |
| Triglycerides | Enter | 0.098 | 0.517 (0.497-0.538) | 97.7% | 2.7% | 0.162 |
| Urate | Enter | 0.105 | 0.517 (0.496-0.538) | 98.9% | 1.3% | 0.113 |
| Urea | Enter | 0.081 | 0.504 (0.484-0.525) | 100% | 0% | 0 |
| Vitamin D | Enter | 0.512 | 0.501 (0.480-0.521) | 100% | 0% | 0 |
| **Immunology** |  |  |  |  |  |  |
| **Red Blood Cell Count** | **Enter** | **0.001** | **0.524 (0.509-0.539)** | **90.7%** | **11.6%** | **0.324** |
| White Blood Cell Count | Enter | 0.426 | 0.505 (0.490-0.520) | 100% | 0.2% | 0.045 |
| C-Reactive Protein | Enter | 0.394 | 0.511 (0.496-0.525) | 99.2% | 0.9% | 0.094 |
| Neutrophils | Enter | 0.071 | 0.521 (0.507-0.536) | 99.4% | 0.7% | 0.083 |
| Lymphocytes | Enter | 0.053 | 0.518 (0.503-0.533) | 99.8% | 0.3% | 0.055 |
| Monocytes | Enter | 0.172 | 0.505 (0.490-0.519) | 98.6% | 1.6% | 0.126 |
| Eosinophils | Enter | 0.853 | 0.514 (0.499-0.528) | 100% | 0% | 0 |
| Basophils | Enter | 0.086 | 0.510 (0.495-0.525) | 100% | 0% | 0 |

Area Under the Curve (AUC); Confidence Interval (CI); Geometric Mean (G-Mean). Specificity and sensitivity are the likelihood of correctly detecting when COVID-19 infection for a test case was negative or positive respectively. G-Mean is the degree to which a given predictor correctly predicts both true negatives
and true positives for COVID-19 infection. “Blue” and “white” shading are used to better visualize predictors within a set of similar variables. P values less than .05 were considered significant and applicable predictors and statistics are bolded.

**Supplementary Table 2.** Isolated effect of each predictor on COVID-19 risk among test cases with serology data.

| Predictor | Classifier Method | P value | AUC (95% CI) | Specificity | Sensitivity | G-Mean |
| --- | --- | --- | --- | --- | --- | --- |
| **Basic Demographics** |  |  |  |  |  |  |
| Age | Enter | 0.634 | 0.568 (0.441-0.694) | 100% | 0% | 0 |
| **Sex** | **Enter** | **<.001** | **0.689 (0.578-0.800)** | **66.0%** | **71.8%** | **0.688** |
| Ethnic Background | Enter | 0.647 | 0.528 (0.407-0.649) | 98.1% | 12.8% | 0.354 |
| Deprivation Index | Enter | 0.704 | 0.539 (0.416-0.662) | 100% | 0% | 0 |
| **Education** | **Enter** | **0.017** | **0.635 (0.522-0.748)** | **64.2%** | **53.8%** | **0.588** |
| **Body Composition** |  |  |  |  |  |  |
| Waist Circumference | Enter | 0.142 | 0.598 (0.480-0.716) | 83.0% | 10.3% | 0.292 |
| Body Mass Index | Enter | 0.575 | 0.548 (0.427-0.669) | 96.2% | 0% | 0 |
| Trunk Fat Mass | Enter | 0.792 | 0.539 (0.413-0.665) | 100% | 0% | 0 |
| Whole Body Fat Mass | Enter | 0.252 | 0.582 (0.464-0.701) | 92.5% | 5.1% | 0.217 |
| **Whole Body Fat-Free Mass** | **Enter** | **0.003** | **0.687 (0.575-0.799)** | **71.7%** | **59.0%** | **0.650** |
| **Whole Body Water Mass** | **Enter** | **0.003** | **0.680 (0.567-0.793)** | **71.7%** | **59.0%** | **0.650** |
| **Health Behaviors and Conditions** |  |  |  |  |  |  |
| Smoking Status | Enter | 0.072 | 0.610 (0.494-0.726) | 47.2% | 0% | 0 |
| Alcohol Status | Enter | 0.094 | 0.603 (0.482-0.723) | 100% | 20.5% | 0.453 |
| Long-Term Medical Condition | Enter | 0.391 | 0.546 (0.427-0.666) | 100% | 0% | 0 |
| Health Rating | Enter | 0.661 | 0.521 (0.403-0.639) | 100% | 0% | 0 |
| **Vitals** |  |  |  |  |  |  |
| Pulse Rate | Enter | 0.335 | 0.582 (0.461-0.702) | 92.5% | 2.6% | 0.155 |
| **Diastolic BP** | **Enter** | **0.047** | **0.633 (0.515-0.752)** | **84.9%** | **35.9%** | **0.552** |
| Systolic BP | Enter | 0.200 | 0.540 (0.420-0.660) | 83.0% | 12.8% | 0.326 |
| **Biochemistry** |  |  |  |  |  |  |
| Alanine Aminotransferase | Enter | 0.303 | 0.588 (0.463-0.714) | 100% | 0% | 0 |
| Albumin | Enter | 0.285 | 0.676 (0.525-0.828) | 93.0% | 0% | 0 |
| Alkaline Phosphatase | Enter | 0.272 | 0.541 (0.422-0.660) | 98.1% | 0% | 0 |
| Apolipoprotein A | Enter | 0.333 | 0.600 (0.463-0.737) | 95.3% | 0% | 0 |
| Apolipoprotein B | Enter | 0.125 | 0.545 (0.426-0.665) | 88.7% | 15.4% | 0.370 |
| Aspartate Aminotransferase | Enter | 0.399 | 0.520 (0.398-0.641) | 96.2% | 0% | 0 |
| Bilirubin (Total) | Enter | 0.319 | 0.543 (0.420-0.665) | 88.7% | 0% | 0 |
| Calcium | Enter | 0.989 | 0.528 (0.386-0.670) | 100% | 0% | 0 |
| Cholesterol (Total) | Enter | 0.276 | 0.528 (0.408-0.649) | 90.6% | 5.1% | 0.215 |
| Creatinine | Enter | 0.250 | 0.524 (0.403-0.646) | 88.7% | 5.1% | 0.213 |
| Cystatin C | Enter | 0.169 | 0.634 (0.512-0.755) | 88.7% | 7.7% | 0.261 |
| Gamma Glutamyltransferase | Enter | 0.103 | 0.604 (0.482-0.725) | 100% | 0% | 0 |
| HDL Cholesterol | Enter | 0.640 | 0.551 (0.410-0.692) | 100% | 0% | 0 |
| **Hemoglobin A1c** | **Enter** | **0.035** | **0.638 (0.523-0.753)** | **90.6%** | **30.8%** | **0.528** |
| Insulin-Like Growth Factor 1 | Enter | 0.852 | 0.533 (0.407-0.659) | 100% | 0% | 0 |
| LDL Cholesterol | Enter | 0.247 | 0.533 (0.413-0.654) | 84.9% | 5.1% | 0.208 |
| Lipoprotein A | Enter | 0.460 | 0.554 (0.418-0.691) | 95.8% | 0% | 0 |
| **Phosphate** | **Enter** | **0.026** | **0.671 (0.531-0.811)** | **88.4%** | **4.3%** | **0.195** |
| **Protein (Total)** | **Enter** | **0.014** | **0.662 (0.514-0.811)** | **90.7%** | **47.8%** | **0.658** |
| Sex Hormone Binding Globulin | Enter | 0.900 | 0.510 (0.360-0.659) | 100% | 0% | 0 |
| **Testosterone** | **Enter** | **0.021** | **0.731 (0.625-0.836)** | **86.8%** | **35.9%** | **0.558** |
| Triglycerides | Enter | 0.063 | 0.614 (0.498-0.729) | 71.7% | 71.8% | 0.717 |
| Urate | Enter | 0.088 | 0.656 (0.538-0.774) | 86.8% | 20.5% | 0.422 |
| **Urea** | **Enter** | **0.021** | **0.632 (0.508-0.757)** | **86.8%** | **35.9%** | **0.558** |
| **Vitamin D** | **Enter** | **0.009** | **0.601 (0.479-0.722)** | **81.1%** | **38.5%** | **0.559** |
| **Immunology** |  |  |  |  |  |  |
| C-Reactive Protein | Enter | 0.394 | 0.567 (0.447-0.687) | 99.2% | 0.9% | 0.094 |
| Red Blood Cell Count | Enter | 0.943 | 0.504 (0.384-0.625) | 90.7% | 11.6% | 0.324 |
| White Blood Cell Count | Enter | 0.426 | 0.592 (0.476-0.709) | 100% | 0.2% | 0.045 |
| **Neutrophils** | **Enter** | **0.037** | **0.628 (0.507-0.749)** | **99.4%** | **0.7%** | **0.083** |
| Lymphocytes | Enter | 0.053 | 0.552 (0.434-0.671) | 99.8% | 0.3% | 0.055 |
| **Monocytes** | **Enter** | **0.020** | **0.646 (0.532-0.761)** | **98.6%** | **1.6%** | **0.126** |
| **Eosinophils** | **Enter** | **0.015** | **0.649 (0.535-0.762)** | **100%** | **0%** | **0** |
| Basophils | Enter | 0.086 | 0.528 (0.409-0.647) | 100% | 0% | 0 |

Area Under the Curve (AUC); Confidence Interval (CI); Geometric Mean (G-Mean). Specificity and sensitivity are the likelihood of correctly detecting when COVID-19 infection for a test case was negative or positive respectively. G-Mean is the degree to which a given predictor correctly predicts both true negatives
and true positives for COVID-19 infection. “Blue” and “white” shading are used to better visualize predictors within a set of similar variables. P values less than .05 were considered significant and applicable predictors and statistics are bolded.

**Supplementary Table 3.** Isolated effect of each baseline antibody titer on predicting current COVID-19 infection risk.

| Pathogen Name | Abbreviation | Antigen | Classifier Method | P value | AUC (95% CI) | Specificity | Sensitivity | G-Mean |
| --- | --- | --- | --- | --- | --- | --- | --- | --- |
| Herpes Simplex Virus-1 | HSV-1 | 1gG | Enter | 0.089 | 0.526 (0.407-0.645) | 56.3% | 51.3% | 0.537 |
| Herpes Simplex Virus-2 | HSV-2 | 2mgG | Enter | 0.422 | 0.518 (0.402-0.634) | 18.8% | 82.1% | 0.393 |
| Varicella Zoster Virus | VZV | gE/gI | Enter | **0.046** | **0.612 (0.509-0.714)** | **31.3%** | **69.2%** | **0.465** |
| Epstein-Barr Virus | EBV | **VCA p18** | **Enter** | **0.002** | **0.675 (0.574-0.775)** | **0%** | **100.0%** | **0** |
|  |  | EBNA-1 | Enter | 0.996 | 0.549 (0.441-0.658) | 0% | 94.9% | 0 |
|  |  | ZEBRA | Enter | 0.419 | 0.589 (0.49-0.688) | 0% | 100.0% | 0 |
|  |  | EA-D | Enter | 0.815 | 0.503 (0.397-0.609) | 0% | 89.7% | 0 |
| Human Cytomegalovirus | CMV | **pp150 Nter** | **Enter** | **0.006** | **0.654 (0.539-0.769)** | **50.0%** | **74.4%** | **0.610** |
|  |  | pp 52 | Enter | 0.285 | 0.555 (0.442-0.668) | 43.8% | 74.4% | 0.571 |
|  |  | pp 28 | Enter | 0.284 | 0.602 (0.485-0.718) | 34.4% | 74.4% | 0.506 |
| Human Herpesvirus-6 | HHV-6 | IE1A | Enter | 0.110 | 0.586 (0.476-0.697) | 31.3% | 76.9% | 0.491 |
|  |  | **IE1B** | **Enter** | **0.007** | **0.638 (0.539-0.738)** | **37.5%** | **82.1%** | **0.555** |
|  |  | p101 k | Enter | 0.687 | 0.532 (0.431-0.633) | 0% | 92.3% | 0 |
| Human Herpesvirus-7 | HHV-7 | **U14** | **Enter** | **0.003** | **0.684 (0.577-0.792)** | **65.6%** | **66.7%** | **0.661** |
| Kaposi's Sarcoma Associated Herpesvirus | KSHV | LANA | Enter | 0.899 | 0.546 (0.439-0.653) | 0% | 94.9% | 0 |
|  |  | K8.1 | Enter | 0.912 | 0.564 (0.452-0.675) | 0% | 94.9% | 0 |
| Hepatitis B Virus | HBV | HBc | Enter | 0.808 | 0.505 (0.399-0.611) | 0% | 100.0% | 0 |
|  |  | HBe | Enter | 0.434 | 0.600 (0.486-0.713) | 3.1% | 82.1% | 0.160 |
| Hepatitis C Virus | HCV | Core | Enter | 0.627 | 0.524 (0.415-0.633) | 0% | 94.9% | 0 |
|  |  | **NS3** | **Enter** | **0.011** | **0.663 (0.559-0.766)** | **50.0%** | **71.8%** | **0.599** |
| Toxoplasma gondii | T. gondii | **p22** | **Enter** | **0.036** | **0.617 (0.517-0.718)** | **40.6%** | **84.6%** | **0.586** |
|  |  | **sag1** | **Enter** | **0.038** | **0.662 (0.554-0.769)** | **37.5%** | **82.1%** | **0.555** |
| Human T Lymphotropic Virus 1 | HTLV-1 | **HTLV-1 gag** | **Enter** | **0.003** | **0.710 (0.618-0.802)** | **56.3%** | **82.1%** | **0.680** |
|  |  | HTLV-1 env | Enter | 0.402 | 0.554 (0.451-0.658) | 18.8% | 92.3% | 0.417 |
|  | HIV | **HIV-1 gag** | **Enter** | **0.001** | **0.688 (0.592-0.785)** | **37.5%** | **84.6%** | **0.563** |
| Human Immunodeficiency Virus |  | HIV-1 env | Enter | 0.204 | 0.577 (0.469-0.685) | 31.3% | 69.2% | 0.465 |
| Human Polyomavirus BKV | BKV | **BK VP1** | **Enter** | **0.008** | **0.649 (0.539-0.759)** | **53.1%** | **59.0%** | **0.560** |
| Human Polyomavirus JCV | JCV | JC VP1 | Enter | 0.796 | 0.530 (0.419-0.641) | 0% | 100.0% | 0 |
| Merkel Cell Polyomavirus | MCV | MC VP1 | Enter | 0.648 | 0.546 (0.442-0.649) | 0% | 89.7% | 0 |
| Human Papillomavirus type-16 | HPV 16 | L1 | Enter | 0.961 | 0.525 (0.412-0.637) | 0% | 100.0% | 0 |
|  |  | **E6** | **Enter** | **0.029** | **0.622 (0.517-0.728)** | **53.1%** | **66.7%** | **0.595** |
|  |  | **E7** | **Enter** | **0.009** | **0.646 (0.536-0.756)** | **34.4%** | **74.4%** | **0.506** |
| Human Papillomavirus type-18 | HPV 18 | L1 | Enter | 0.404 | 0.556 (0.453-0.660) | 12.5% | 84.6% | 0.325 |
| Chlamydia trachomatis | C. trachomatis | momp D | Enter | 0.455 | 0.501 (0.390-0.612) | 3.1% | 97.4% | 0.174 |
|  |  | momp A | Enter | 0.075 | 0.551 (0.450-0.653) | 28.1% | 89.7% | 0.502 |
|  |  | tarp-D F1 | Enter | 0.918 | 0.549 (0.431-0.668) | 0% | 100.0% | 0 |
|  |  | tarp-D F2 | Enter | 0.814 | 0.572 (0.459-0.686) | 0% | 100.0% | 0 |
|  |  | PorB | Enter | 0.352 | 0.597 (0.488-0.706) | 9.4% | 87.2% | 0.286 |
|  |  | **pGP3** | **Enter** | **0.005** | **0.656 (0.547-0.765)** | **21.9%** | **79.5%** | **0.417** |
| Helicobacter pylori | H. pylori | CagA* | N/A | N/A | N/A | N/A | N/A | N/A |
|  |  | **VacA** | **Enter** | **0.045** | **0.613 (0.506-0.719)** | **25.0%** | **87.2%** | **0.467** |
|  |  | OMP | Enter | 0.770 | 0.509 (0.397-0.622) | 0% | 94.9% | 0 |
|  |  | GroEL | Enter | 0.308 | 0.591 (0.467-0.715) | 28.1% | 74.4% | 0.457 |
|  |  | Catalase | Enter | 0.663 | 0.525 (0.416-0.634) | 0% | 89.7% | 0 |
|  |  | UreA | Enter | 0.290 | 0.567 (0.461-0.672) | 18.8% | 87.2% | 0.405 |

Area Under the Curve (AUC); Confidence Interval (CI); Geometric Mean (G-Mean). Specificity and sensitivity are the likelihood of correctly detecting if COVID-19 infection for a test case was negative or positive respectively. G-Mean is the degree to which a given antigen correctly predicts both true negatives and true positives for COVID-19 infection. “Blue” and “white” shading are used to better visualize antigens specific to a given pathogen. P values less than .05 were considered significant and applicable antigens and statistics are bolded. \*The CagA antigen was excluded from analysis due to roughly half of sample analyte values being lost to lab error.

**Supplementary Table 4.**
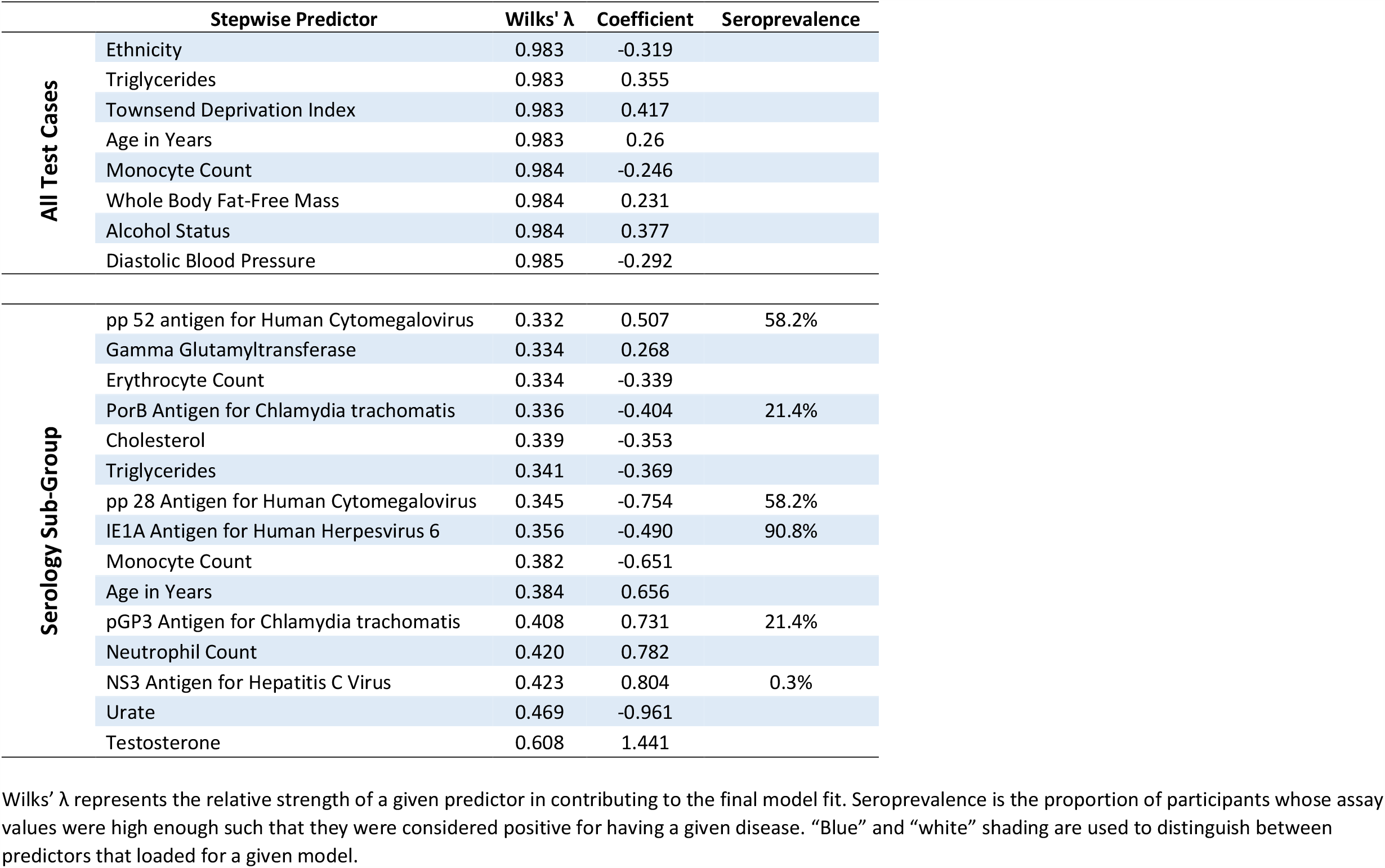
Predictors that loaded into the stepwise models for COVID-19 infection risk.

| | Stepwise Predictor | Wilks' $\lambda$ | Coefficient | Seroprevalence |
| --- | --- | --- | --- | --- |
| All Test Cases | Ethnicity | 0.983 | -0.319 |  |
|  | Triglycerides | 0.983 | 0.355 |  |
|  | Townsend Deprivation Index | 0.983 | 0.417 |  |
|  | Age in Years | 0.983 | 0.26 |  |
|  | Monocyte Count | 0.984 | -0.246 |  |
|  | Whole Body Fat-Free Mass | 0.984 | 0.231 |  |
|  | Alcohol Status | 0.984 | 0.377 |  |
|  | Diastolic Blood Pressure | 0.985 | -0.292 |  |
| Serology Sub-Group | pp 52 antigen for Human Cytomegalovirus | 0.332 | 0.507 | 58.2% |
|  | Gamma Glutamyltransferase | 0.334 | 0.268 |  |
|  | Erythrocyte Count | 0.334 | -0.339 |  |
|  | PorB Antigen for Chlamydia trachomatis | 0.336 | -0.404 | 21.4% |
|  | Cholesterol | 0.339 | -0.353 |  |
|  | Triglycerides | 0.341 | -0.369 |  |
|  | pp 28 Antigen for Human Cytomegalovirus | 0.345 | -0.754 | 58.2% |
|  | IE1A Antigen for Human Herpesvirus 6 | 0.356 | -0.490 | 90.8% |
|  | Monocyte Count | 0.382 | -0.651 |  |
|  | Age in Years | 0.384 | 0.656 |  |
|  | pGP3 Antigen for Chlamydia trachomatis | 0.408 | 0.731 | 21.4% |
|  | Neutrophil Count | 0.420 | 0.782 |  |
|  | NS3 Antigen for Hepatitis C Virus | 0.423 | 0.804 | 0.3% |
|  | Urate | 0.469 | -0.961 |  |
|  | Testosterone | 0.608 | 1.441 |  |

Wilks' $\lambda$ represents the relative strength of a given predictor in contributing to the final model fit. Seroprevalence is the proportion of participants whose assay values were high enough such that they were considered positive for having a given disease. "Blue" and "white" shading are used to distinguish between predictors that loaded for a given model.

**Supplementary Table 5.** Isolated effect of each predictor on COVID-19 severity among the full sample.

| Predictor | Classifier Method | P value | AUC (95% CI) | Specificity | Sensitivity | G-Mean |
| --- | --- | --- | --- | --- | --- | --- |
| **Basic Demographics** |  |  |  |  |  |  |
| Age | Enter | <.001 | **0.572 (0.548-0.596)** | **75.5%** | **36.7%** | **0.526** |
| Sex | Enter | **0.009** | **0.528 (0.504-0.552)** | **100%** | **0%** | **0** |
| Ethnic Background | Enter | <.001 | **0.524 (0.500-0.548)** | **91.0%** | **12.9%** | **0.343** |
| Deprivation Index | Enter | 0.610 | 0.507 (0.483-0.531) | 91.0% | 12.9% | 0.343 |
| Education | Enter | 0.854 | 0.505 (0.478-0.532) | 100% | 0% | 0 |
| **Body Composition** |  |  |  |  |  |  |
| Waist Circumference | Enter | **0.003** | **0.541 (0.517-0.565)** | **93.6%** | **6.0%** | **0.237** |
| Body Mass Index | Enter | 0.198 | 0.522 (0.498-0.547) | 100% | 0% | 0 |
| Trunk Fat Mass | Enter | 0.068 | 0.531 (0.506-0.555) | 99.9% | 0.1% | 0.032 |
| Whole Body Fat Mass | Enter | 0.341 | 0.521 (0.497-0.546) | 100% | 0% | 0 |
| Whole Body Fat-Free Mass | Enter | 0.104 | 0.522 (0.498-0.547) | 100% | 0% | 0 |
| Whole Body Water Mass | Enter | 0.095 | 0.522 (0.497-0.546) | 100% | 0% | 0 |
| **Health Behaviors and Conditions** |  |  |  |  |  |  |
| Smoking Status | Enter | 0.114 | 0.522 (0.497-0.546) | 100% | 0% | 0 |
| Alcohol Status | Enter | 0.540 | 0.506 (0.482-0.530) | 100% | 0% | 0 |
| Long-Term Medical Condition | Enter | 0.084 | 0.519 (0.494-0.543) | 100% | 0% | 0 |
| Health Rating | Enter | 0.098 | 0.518 (0.494-0.543) | 100% | 0% | 0 |
| **Vitals** |  |  |  |  |  |  |
| Pulse Rate | Enter | 0.652 | 0.510 (0.486-0.535) | 100% | 0% | 0 |
| Diastolic BP | Enter | 0.969 | 0.501 (0.476-0.526) | 100% | 0% | 0 |
| Systolic BP | Enter | **0.008** | **0.540 (0.515-0.565)** | **98.0%** | **2.0%** | **0.140** |
| **Biochemistry** |  |  |  |  |  |  |
| Alanine Aminotransferase | Enter | **0.039** | **0.540 (0.515-0.565)** | **100%** | **0%** | **0** |
| Albumin | Enter | 0.093 | 0.525 (0.498-0.552) | 98.9% | 0.4% | 0.063 |
| Alkaline Phosphatase | Enter | 0.198 | 0.522 (0.496-0.547) | 100% | 0% | 0 |
| Apolipoprotein A | Enter | 0.245 | 0.513 (0.487-0.540) | 99.7% | 0.1% | 0.032 |
| Apolipoprotein B | Enter | 0.862 | 0.502 (0.477-0.528) | 100% | 0% | 0 |
| **Aspartate Aminotransferase** | **Enter** | **0.047** | **0.534 (0.508-0.559)** | **100%** | **0%** | **0** |
| Bilirubin (Total) | Enter | 0.965 | 0.509 (0.483-0.534) | 100% | 0% | 0 |
| Calcium | Enter | 0.545 | 0.507 (0.481-0.534) | 99.8% | 0% | 0 |
| Cholesterol (Total) | Enter | 0.352 | 0.510 (0.484-0.535) | 100% | 0% | 0 |
| Creatinine | Enter | 0.289 | 0.510 (0.485-0.535) | 100% | 0% | 0 |
| **Cystatin C** | **Enter** | **0.006** | **0.527 (0.502-0.553)** | **99.5%** | **0.7%** | **0.083** |
| Gamma Glutamyltransferase | Enter | 0.181 | 0.529 (0.504-0.555) | 100% | 0% | 0 |
| HDL Cholesterol | Enter | 0.180 | 0.517 (0.490-0.544) | 98.7% | 0.4% | 0.063 |
| **Hemoglobin A1c** | **Enter** | **0.002** | **0.555 (0.529-0.580)** | **99.3%** | **0.7%** | **0.083** |
| **Insulin-Like Growth Factor 1** | **Enter** | **0.037** | **0.524 (0.499-0.550)** | **96.8%** | **3.8%** | **0.192** |
| LDL Cholesterol | Enter | 0.470 | 0.508 (0.483-0.533) | 100% | 0% | 0 |
| Lipoprotein A | Enter | 0.216 | 0.518 (0.489-0.546) | 100% | 0% | 0 |
| Phosphate | Enter | 0.357 | 0.513 (0.486-0.540) | 99.9% | 0% | 0 |
| Protein (Total) | Enter | 0.930 | 0.512 (0.486-0.539) | 100% | 0% | 0 |
| **Sex Hormone Binding Globulin** | **Enter** | **0.036** | **0.525 (0.498-0.552)** | **94.2%** | **7.5%** | **0.266** |
| Testosterone | Enter | 0.060 | 0.521 (0.495-0.548) | 100% | 0% | 0 |
| Triglycerides | Enter | 0.060 | 0.528 (0.503-0.554) | 100% | 0% | 0 |
| **Urate** | **Enter** | **0.012** | **0.533 (0.508-0.559)** | **98.5%** | **1.7%** | **0.129** |
| **Urea** | **Enter** | **0.003** | **0.530 (0.505-0.556)** | **97.8%** | **2.7%** | **0.162** |
| Vitamin D | Enter | 0.562 | 0.503 (0.477-0.529) | 100% | 0% | 0 |
| **Immunology** |  |  |  |  |  |  |
| Red Blood Cell Count | Enter | 0.732 | 0.504 (0.479-0.529) | 100% | 0% | 0 |
| **White Blood Cell Count** | **Enter** | **0.025** | **0.536 (0.511-0.561)** | **100%** | **0%** | **0** |
| C-Reactive Protein | Enter | 0.598 | 0.528 (0.503-0.554) | 100% | 0% | 0 |
| **Neutrophils** | **Enter** | **0.004** | **0.535 (0.510-0.560)** | **98.2%** | **3.6%** | **0.188** |
| Lymphocytes | Enter | 0.212 | 0.504 (0.479-0.530) | 100% | 0% | 0 |
| Monocytes | Enter | 0.071 | 0.530 (0.505-0.555) | 100% | 0% | 0 |
| Eosinophils | Enter | 0.291 | 0.520 (0.495-0.545) | 100% | 0% | 0 |
| Basophils | Enter | 0.671 | 0.500 (0.475-0.525) | 100% | 0% | 0 |

Area Under the Curve (AUC); Confidence Interval (CI); Geometric Mean (G-Mean). Here, specificity and sensitivity are the likelihood of correctly detecting if a
positive COVID-19 test case was mild or severe respectively. G-Mean is the degree to which a given predictor correctly predicts both true negatives and true positives for COVID-19 infection severity. "Orange" and "white" shading are used to better visualize each class of predictors for COVID-19 severity. P values less than .05 were considered significant, where applicable predictors and classifier statistics are bolded.

**Supplementary Table 6.**
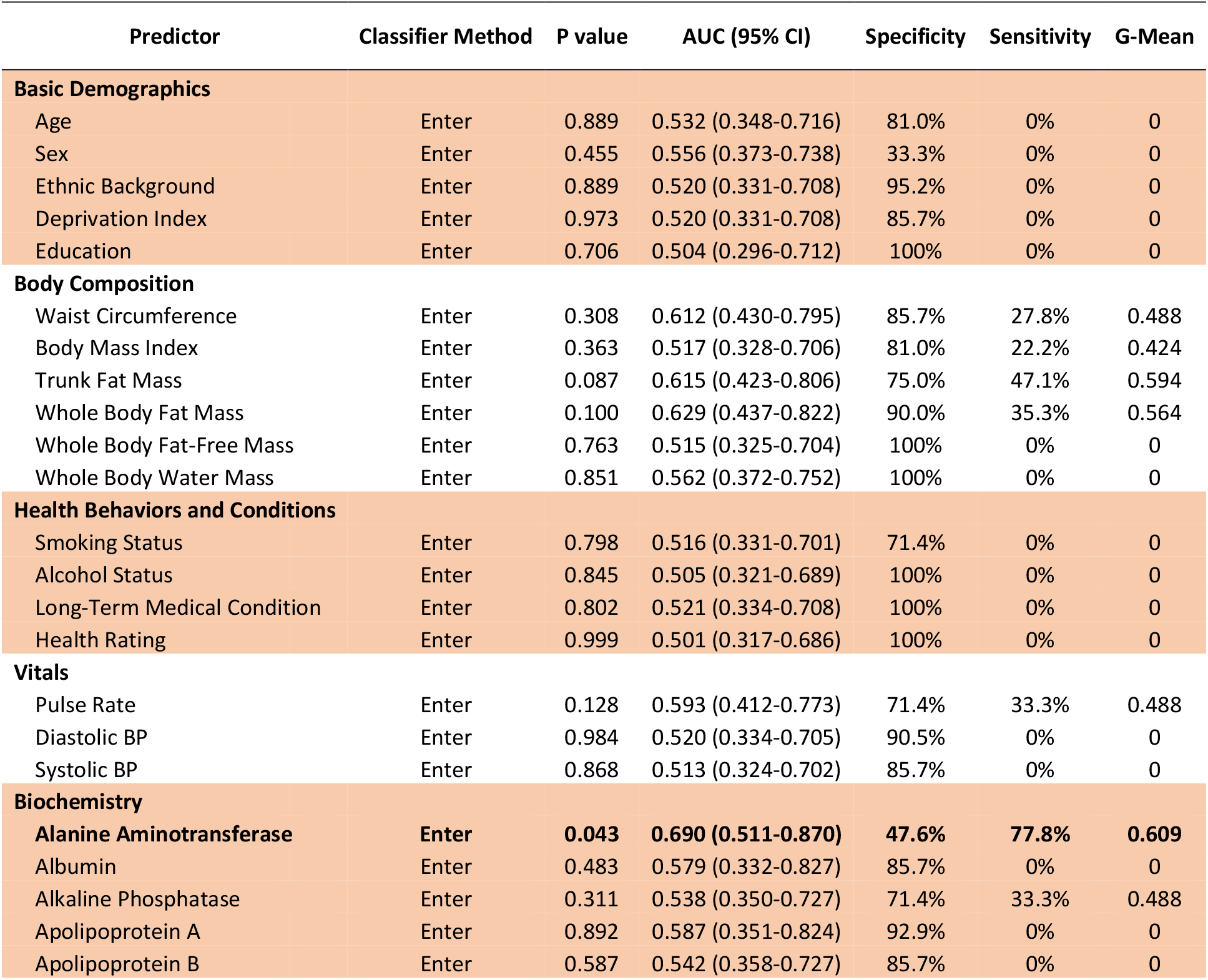
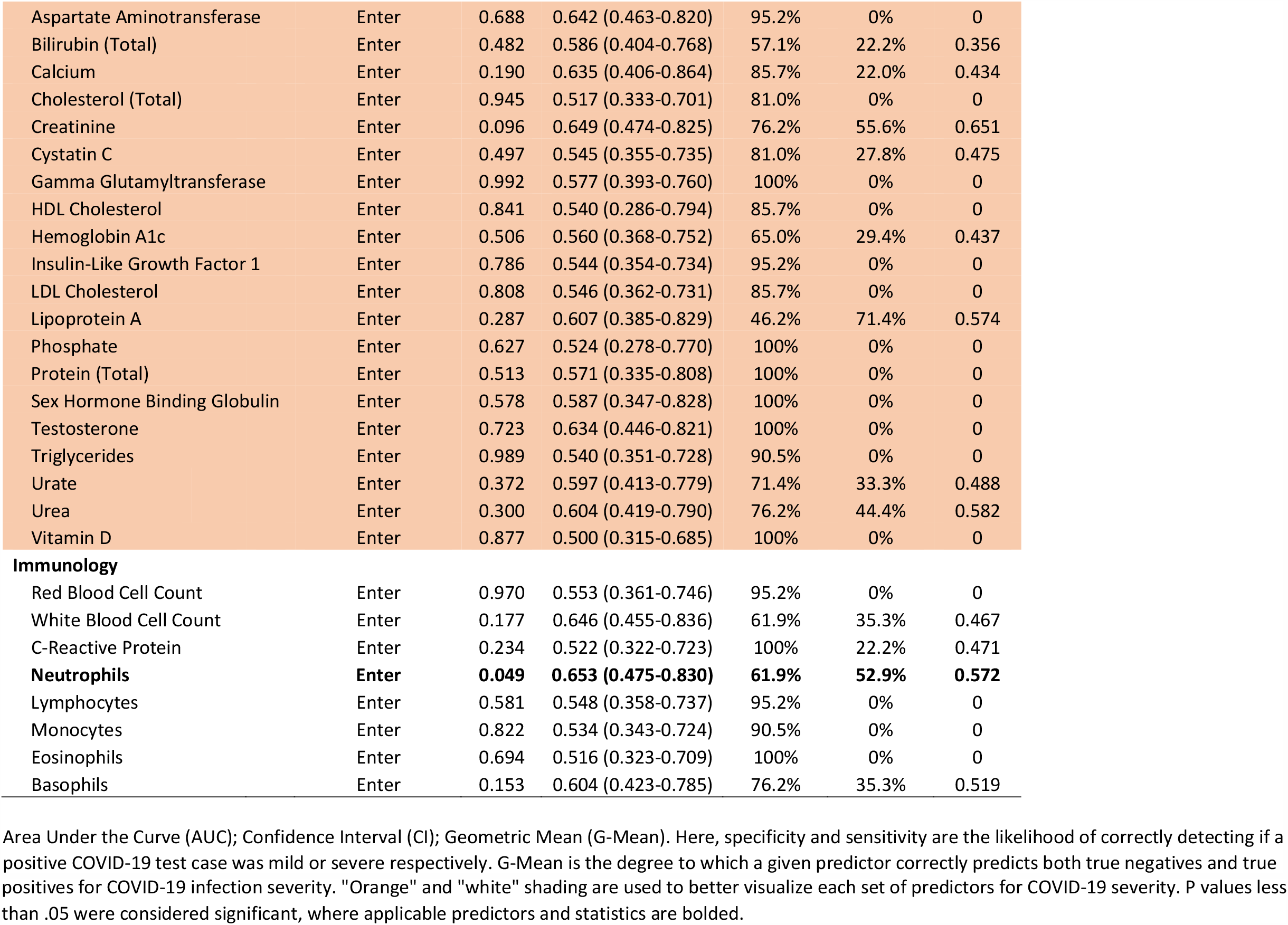
Isolated effect of each predictor on COVID-19 severity for the serology sub-group.

**Supplementary Table 7.**
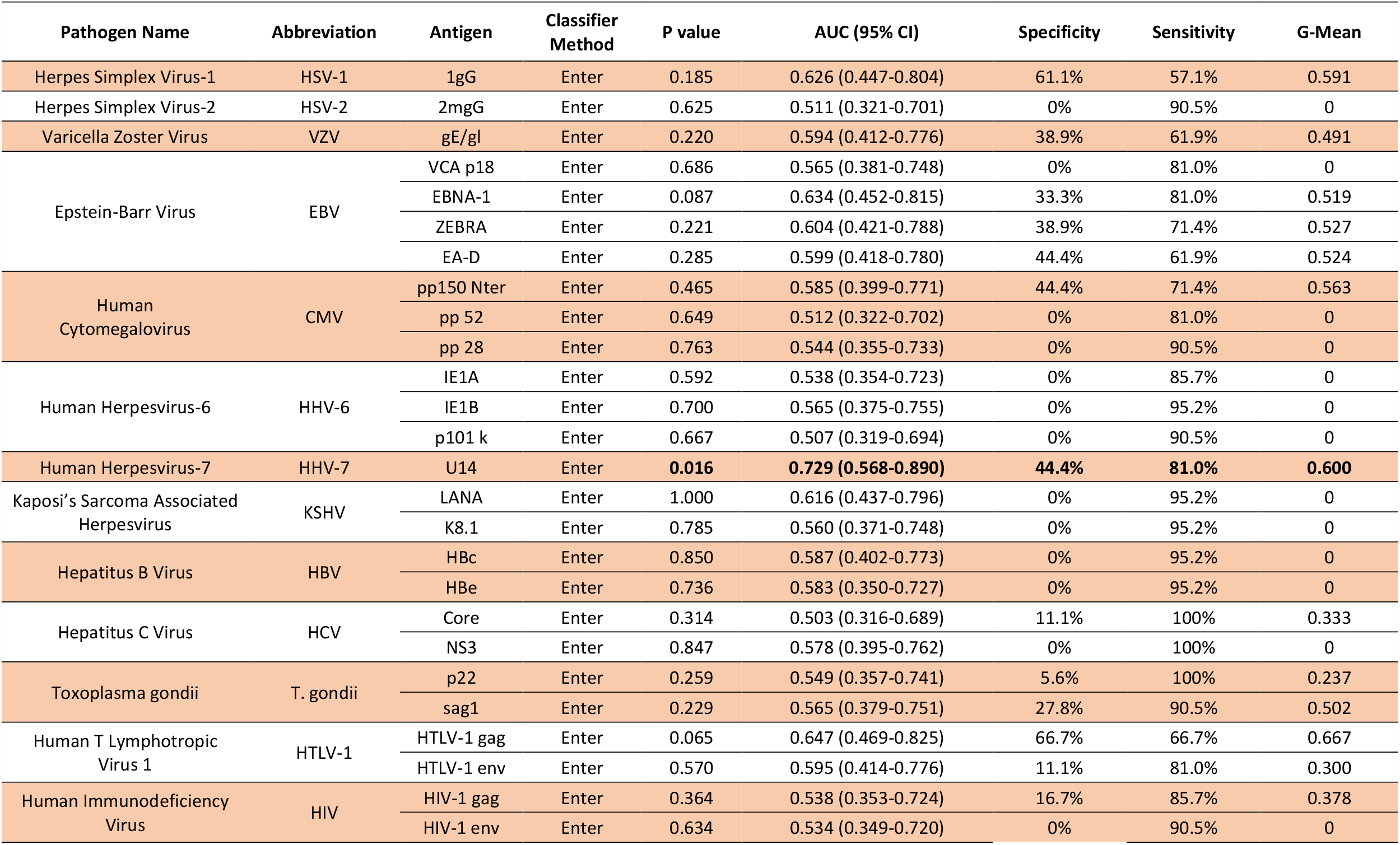
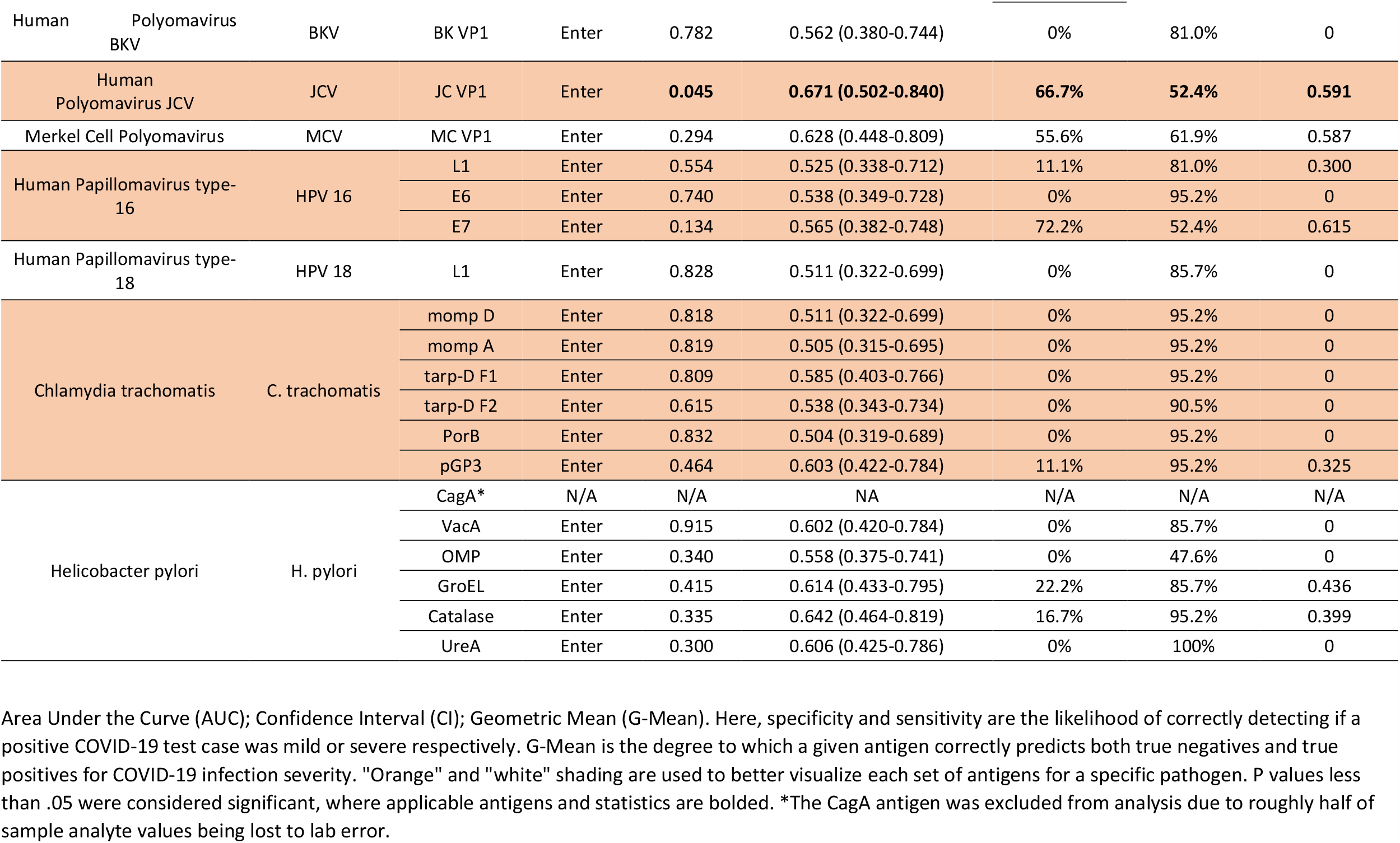
Isolated effect of each baseline antibody titer on predicting current COVID-19 infection severity.

**Supplementary Table 8.**
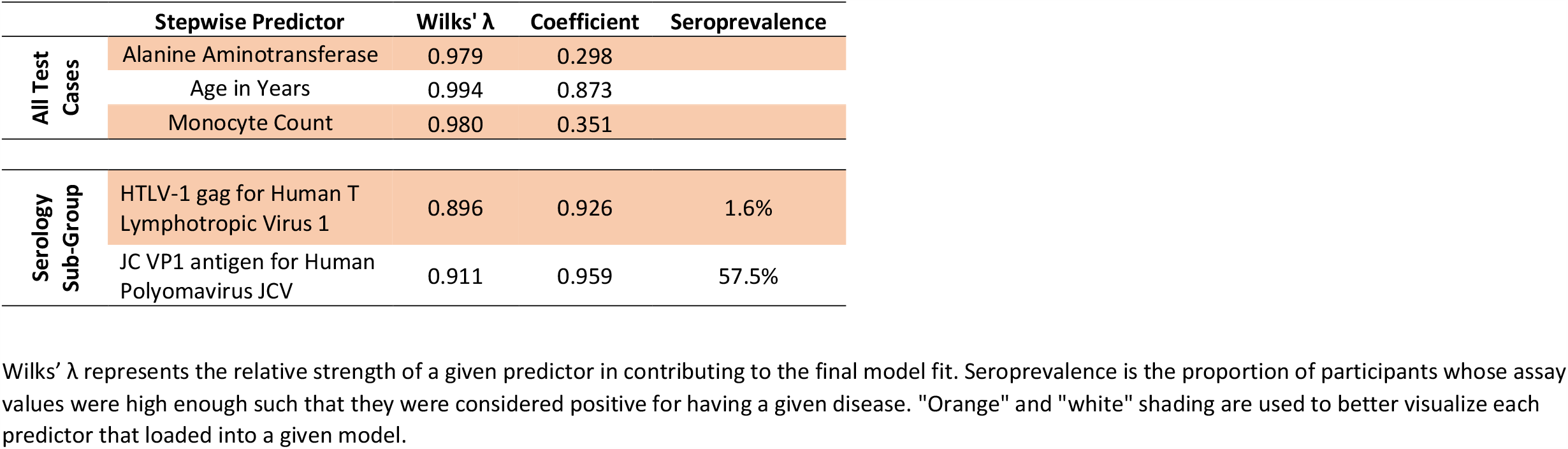
Predictors that loaded into the stepwise models for COVID-19 severity risk.

## References

1 Coronaviridae Study Group of the International Committee on Taxonomy of, V. The species Severe acute respiratory syndrome-related coronavirus: classifying 2019-nCoV and naming it SARS-CoV-2. Nat Microbiol 5, 536–544, doi:10.1038/s41564-020-0695-z (2020).

2 Sattar, N., McInnes, I. B. & McMurray, J. J. V. Obesity a Risk Factor for Severe COVID-19 Infection: Multiple Potential Mechanisms. Circulation, doi:10.1161/CIRCULATIONAHA.120.047659 (2020).

3 Simonnet, A. et al. High prevalence of obesity in severe acute respiratory syndrome coronavirus-2 (SARS-CoV-2) requiring invasive mechanical ventilation. Obesity (Silver Spring), (doi:10.1002/oby.22831 2020).

4 Zhou, F. et al. Clinical course and risk factors for mortality of adult inpatients with COVID-19 in Wuhan, China: a retrospective cohort study. Lancet 395, 1054–1062, doi:10.1016/S0140-6736(20)30566-3 (2020).

5 Patel, A. P., Paranjpe, M. D., Kathiresan, N. P., Rivas, M. A. & Khera, A.V. Race, Socioeconomic Deprivation, and Hospitalization for COVID-19 in English participants of a National Biobank. medRxiv, 2020.2004.2027.20082107, doi:10.1101/2020.04.27.20082107 (2020).

6 Hamer, M., Kivimaki, M., Gale, C. R. & David Batty, G. Lifestyle risk factors, inflammatory mechanisms, and COVID-19 hospitalization: A community-based cohort study of 387,109 adults in UK. Brain Behav Immun, doi:10.1016/j.bbi.2020.05.059 (2020).

7 Liu, Y. et al. Viral dynamics in mild and severe cases of COVID-19. Lancet Infect Dis 20, 656–657, doi:10.1016/S1473-3099(20)30232-2 (2020).

8 Qin, C. et al. Dysregulation of immune response in patients with COVID-19 in Wuhan, China. Clin Infect Dis, doi:10.1093/cid/ciaa248 (2020).

9 Li, T. et al. Significant changes of peripheral T lymphocyte subsets in patients with severe acute respiratory syndrome. J Infect Dis 189, 648–651, doi:10.1086/381535 (2004).

10 Moss, P. ‘The ancient and the new’: is there an interaction between cytomegalovirus and SARS-CoV-2 infection? Immun Ageing 17, 14, doi:10.1186/s12979-020-00185-x (2020).

11 Chidrawar, S. et al. Cytomegalovirus-seropositivity has a profound influence on the magnitude of major lymphoid subsets within healthy individuals. Clin Exp Immunol 155, 423–432, doi:10.1111/j.1365-2249.2008.03785.x (2009).

12 Sudlow, C. et al. UK Biobank: An open access resource for identifying the causes of a wide range of complex diseases of middle and old age. PLOS Medicine 12, e1001779, doi:10.1371/journal.pmed.1001779 (2015).

13 Armstrong, J. et al. Dynamic linkage of COVID-19 test results between Public Health England’s Second Generation Surveillance System and UK Biobank.[Google Scholar]. (2020).

14 Hilton, B. et al. Incidence of Microbial Infections in English UK Biobank Participants: Comparison with the General Population. medRxiv, 2020.2003.2018.20038281, doi:10.1101/2020.03.18.20038281 (2020).

15 Phillimore, P., Beattie, A. & Townsend, P. Widening inequality of health in northern England, 1981-91. Bmj 308, 1125–1128 (1994).

16 Klinedinst, B. S. et al. Aging-related changes in fluid intelligence, muscle and adipose mass, and sex-specific immunologic mediation: A longitudinal UK Biobank study. Brain Behav Immun 82, 396–405, doi:10.1016/j.bbi.2019.09.008 (2019).

17 Kotler, D. P., Burastero, S., Wang, J. & Pierson, R. N., Jr. Prediction of body cell mass, fat-free mass, and total body water with bioelectrical impedance analysis: effects of race, sex, and disease. Am J Clin Nutr 64, 489S–497S, doi:10.1093/ajcn/64.3.489S (1996).

18 Elliott, P. & Peakman, T. C. The UK Biobank sample handling and storage protocol for the collection, processing and archiving of human blood and urine. Int J Epidemiol 37, 234–244, doi:10.1093/ije/dym276 (2008).

19 Waterboer, T., Sehr, P. & Pawlita, M. Suppression of non-specific binding in serological Luminex assays. J Immunol Methods 309, 200–204, doi:10.1016/j.jim.2005.11.008 (2006).

20 Hastie, T., Tibshirani, R. & Friedman, J. The elements of statistical learning: data mining, inference, and prediction. (Springer Science & Business Media, 2009).

21 Marron, J. S., Todd, M. J. & Ahn, J. Distance-weighted discrimination. Journal of the American Statistical Association 102, 1267–1271 (2007).

22 Mundry, R. & Sommer, C. Discriminant function analysis with nonindependent data: consequences and an alternative. Animal Behaviour 74, 965–976 (2007).

23 Hair Jr, J. F., Anderson, R. E., Tatham, R. L. & Black, C. Multivariate data analysis with readings. (Prentice Hall, 1995).

24 Efron, B. in Breakthroughs in statistics 569–593 (Springer, 1992).

25 Weinberger, B. et al. Healthy aging and latent infection with CMV lead to distinct changes in CD8+ and CD4+ T-cell subsets in the elderly. Hum Immunol 68, 86–90, doi:10.1016/j.humimm.2006.10.019 (2007).

26 Osborn, J. E. et al. Comparison of JC and BK human papovaviruses with simian virus 40: restriction endonuclease digestion and gel electrophoresis of resultant fragments. Journal of Virology 13, 614–622 (1974).

27 Liu, W. et al. Analysis of factors associated with disease outcomes in hospitalized patients with 2019 novel coronavirus disease. Chin Med J (Engl) 133, 1032–1038, doi:10.1097/CM9.0000000000000775 (2020).

28 Wang, D. et al. Clinical characteristics of 138 hospitalized patients with 2019 novel coronavirus–infected pneumonia in Wuhan, China. Jama 323, 1061–1069 (2020).

29 Maggio, M. et al. The relationship between testosterone and molecular markers of inflammation in older men. J Endocrinol Invest 28, 116–119 (2005).

30 Qiao, Z., Zhou, L. & Huang, J. Z. Sparse Linear Discriminant Analysis with Applications to High Dimensional Low Sample Size Data. International Journal of Applied Mathematics 39 (2009).

31 Rausch, J. R. & Kelley, K. A comparison of linear and mixture models for discriminant analysis under nonnormality. Behavior Research Methods 41, 85–98 (2009).

32 Pohar, M., Blas, M. & Turk, S. Comparison of logistic regression and linear discriminant analysis: a simulation study. Metodoloski zvezki 1, 143 (2004).

33 Ye, J. et al. in Proceedings of the 15th ACM international conference on Information and knowledge management. 532–539.

